# Silencer variants are key drivers of gene upregulation in Alzheimer’s disease

**DOI:** 10.1101/2025.04.07.25325386

**Authors:** Di Huang, Ivan Ovcharenko

**Affiliations:** Division of Intramural Research, National Library of Medicine, National Institutes of Health, Bethesda, MD, 20892, USA

## Abstract

Alzheimer’s disease (AD), particularly late-onset AD, stands as the most prevalent neurodegenerative disorder globally. Owing to its substantial heritability, genetic studies have emerged as indispensable for elucidating genes and biological pathways driving AD onset and progression. However, genetic and molecular mechanisms underlying AD remain poorly defined, largely due to the pronounced heterogeneity of AD and the intricate interactions among AD genetic factors. Notably, approximately 90% of AD-associated genetic variants reside in intronic and intergenic regions, yet their functional significance has remained largely uncharacterized.

To address this challenge, we developed a deep learning framework combining bulk and single-cell epigenomic data to evaluate the regulatory potential (i.e., silencing and activating strength) of noncoding AD variants in the dorsolateral prefrontal cortex (DLPFCs) and its major cell types. This model identified 1,457 silencer and 3,084 enhancer AD-associated variants in the DLPFC and binned them into silencer variants only (SL), enhancer variants only (EN), or both variant types (ENSL) classes. Each class exerts distinct cellular and molecular influences on AD pathogenesis. EN loci predominantly regulate housekeeping metabolic processes, whereas SL loci (including the genes *MS4A6A*, *TREM2*, *USP6NL*, *HLA-D*) are selectively linked to immune responses. Notably, 71% of these genes are significantly upregulated in AD and pro-inflammation-stimulated microglia. Furthermore, genes associated with SL loci are, in neuronal cells, often responsive to glutamate receptor antagonists (e.g, NBQX) and anti-inflammatory perturbagens (such as D-64131), the compound classes known for reducing the AD risk. ENSL loci, in contrast, are uniquely implicated in memory maintenance, neurofibrillary tangle assembly, and are also shared by other neurological disorders such as Parkinson’s disease and schizophrenia. Key genes in this class of loci, such as *MAPT*, *CR1/2*, and *CLU*, are frequently upregulated in AD subtypes with hyperphosphorylated tau aggregates.

Critically, our model can accurately predict the impact of regulatory variants, with an average Pearson correlation coefficient of 0.54 and a directional concordance rate of 70% between our predictions and experimental outcomes. This model identified rs636317 as a causal AD variant in the *MS4A* locus, distinguishing it from the 7bp-away allele-neutral variant rs636341. Similarly, rs7922621 was prioritized over its 54-bp-away allele-neutral rs7901634 in the *TSPAN14* locus. Additional causal variants include rs6701713 in the *CR1* locus, and rs28834970 and rs755951 in the *PTK2B* locus. Collectively, this work advances our understanding of the regulatory landscape of AD-associated genetic variants, providing a framework to explore their functional roles in the pathogenesis of this complex disease.

## Introduction

Alzheimer’s disease (AD) is the most common neurodegenerative disorder among the elderly, representing a rapidly escalating global epidemic ^1,2^. It is projected that, by 2060, 13.8 million Americans will be affected by AD, imposing substantial societal and economic burdens ^3^. Pathologically, AD is characterized by two hallmark features – the accumulation of amyloid beta (Aβ) into extracellular plaques and the aggregation of hyperphosphorylated tau into neurofibrillary tangles within neurons ^4^. Beyond these features, AD brains exhibit profound dysregulation of immune responses, impaired glucose and lipid metabolism, and vulnerable brain vasculature, among other abnormalities ^5–8^. The etiology of AD is profoundly complex and multifactorial, posing great research challenges ^7,9^.

Due to the high genetic heritability of AD (58 to 79%, varying across investigation contexts) ^10^, genetics analysis has emerged as a powerful tool for unraveling its underlying mechanisms ^11^. These approaches have identified pivotal AD-associated genes and biological pathways. However, the precise molecular underpinnings of AD remain incompletely understood. In particular, most AD-associated variants reside in intronic or intergenic regions, and their regulatory roles remain largely unexplored ^12^. To bridge this knowledge gap, multi-omics approaches, such as genome-wide transcriptomic and epigenomic data, have been leveraged to elucidate how these non-coding variants influence gene expression in AD brains ^13–15^. Recent advances in single-cell/nucleus sequencing techniques (scRNA-seq/snRNA-seq and scATAC-seq) have provided critical insights into cell-type-specific contributions to AD pathological burdens ^12,16–18^. For example, transcriptomics analysis for cerebrovascular cells has linked brain-blood barrier breakdown with *APOE4*-dependent cognitive decline ^16^. Neurons, especially excitatory neurons, experience substantial losses in chromatin accessibility in AD brains ^12^. Despite these advances, existing single-cell epigenomic studies largely focus on chromatin accessibility, leaving the directionality (activating vs repressing) of regulatory effects unexplored.

Furthermore, massively parallel reporter assays (MPRAs) have been utilized to simultaneously assess the regulatory influence of up to tens of thousands of non-coding variants. While these assays have identified variants with significant effects on gene expression ^19–22^, most MPRA experiments have been conducted in immortalized cell lines, such as HEK293 and K562, raising concerns about the relevance of MPRA findings to *in vivo* conditions.

Here, to assess the functional roles of intronic and intergenic variants in the dorsolateral prefrontal cortex (DLPFC), we developed a deep learning model that integrates large-scale complementary epigenomic profiles of bulk and single-cell levels. This approach identifies thousands of enhancer and silencer variants among over 18,000 single nucleotide variants (SNVs) associated with AD in genome-wide association studies (GWASs). The distribution of these variants classifies AD susceptibility loci into three distinct classes. Each class, associated with unique transcriptomic and epigenomic patterns in the healthy and AD DLPFCs, exhibits specific molecular and cellular functions during AD progression. Furthermore, by prioritizing candidate causal regulatory variants for AD, this study sheds light on the regulatory mechanisms underlying AD pathogenesis.

## Results

### Deep learning profiles AD-associated regulatory variants in the DLPFC

For a comprehensive investigation, we compiled AD-associated variants from three recent GWAS studies, each involving hundreds of thousands to millions of participants ^14,23,24^. In total, we collected 18,826 SNVs significantly associated with AD in at least one of these studies and referred to them as adSNVs (Figure 1A). Consistent with prior observations, 91.1% of adSNVs reside in intronic or intergenic regions, categorized as distal-regulatory-element (distal-RE) variants. By merging adjacent gene loci containing adSNVs, we defined 124 distinct AD susceptibility loci (see Methods, Table S1). Among them, 24 (19%) are enriched for promoter/exon adSNVs, while 49 loci (40%) predominantly harbor distal-RE variants, with over 95% of adSNVs residing within distal REs (Figure 1B). These observations underscore the prominent roles of distal-RE adSNVs in AD pathogenesis, emphasizing the imperative to decipher their regulatory functions.

**Figure 1.**
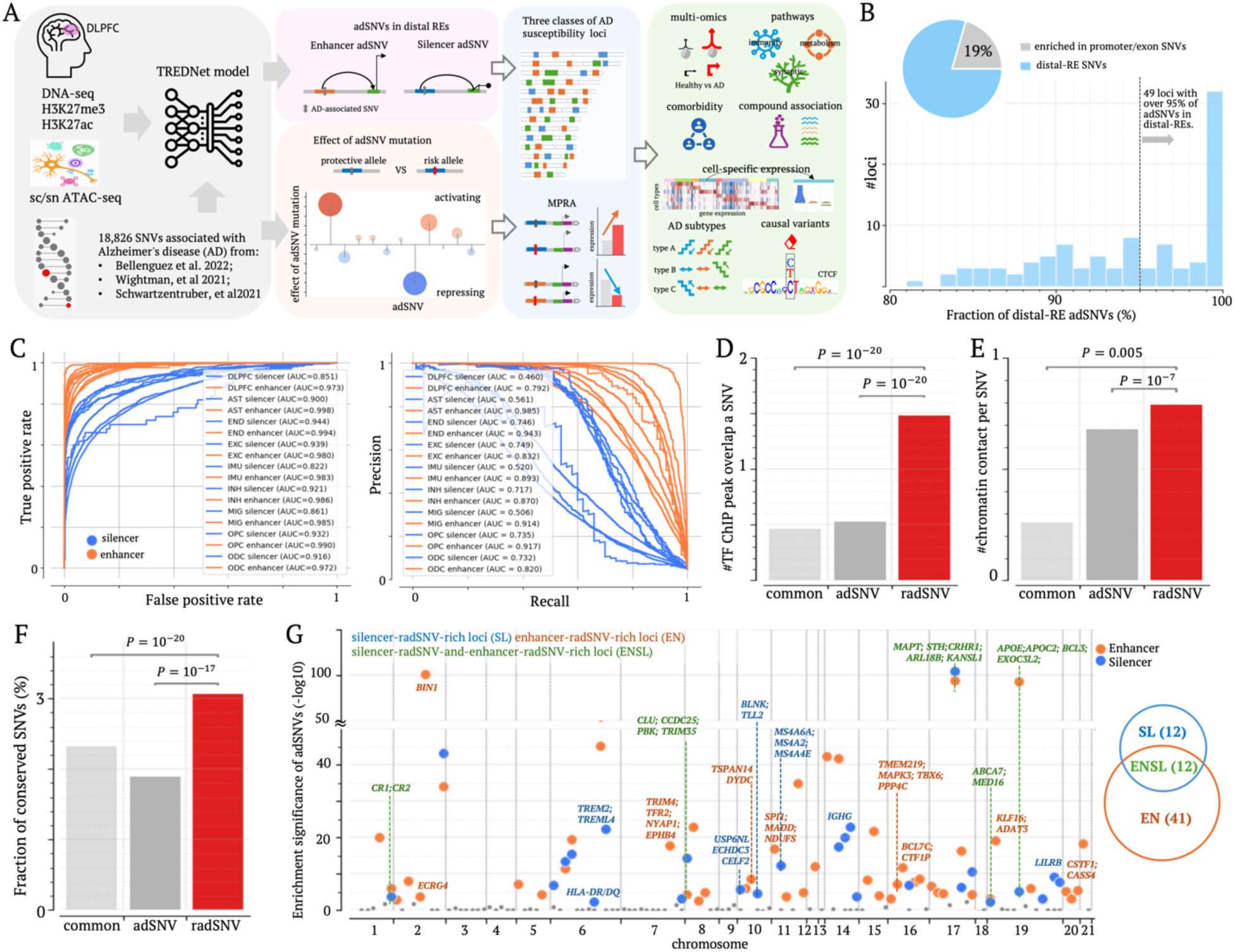
Profiling silencer and enhancer adSNVs with TREDNet. (A) Schematics of the analysis workflow for the identification of radSNVs in the DLPFC and subsequent analyses. (B) Distribution of adSNVs in distal-REs and exon/promoter regions within individual AD susceptibility loci. (C) Classification performance (auROCs and auPRCs) of TREDNet models for silencers and enhancers in the DLPFC and its cell types. (D) Overlap of TF ChIP-seq peaks per SNV across SNV groups. (E) chromatin contact frequencies across SNV groups. (F) Proportion of SNVs located within evolutionarily conserved regions. (G) Genomic distribution plot for AD susceptibility loci. Different locus classes are represented by different colors.

To annotate distal-RE adSNVs, we adapted a two-phase deep learning model TREDNet ^25^ to predict enhancers and silencers in the human DLPFC and its major cell types, including excitatory and inhibitory neurons, astrocytes, endothelial cells, microglia, oligodendrocytes, oligodendrocyte progenitor cells, and immune cells. For training, we compiled DNase I hypersensitive site sequencing peaks (DNase-seq) and chromatin immunoprecipitation sequencing peaks (ChIP-seq) for histone modifications H3K27ac and H3K27me3 from postmortem brain samples of 20 elderly undemented cases (the average age at death was 89 years) ^26^. ScATAC-seq or snATAC-seq data for brain tissues from three recent studies ^17,27,28^ were also incorporated in the TREDNet. Using a multi-task cost function, the TREDNet model was optimized to predict enhancers and silencers in the DLPFC and its cell types (see Methods). The resulting model exhibited robust performance in cross-validation, achieving an area under the receiver operating characteristic curve (auROC) of 0.985 for enhancers and 0.899 for silencers, and an area under the precision-recall curve (auPRC) of 0.885 for enhancers and 0.637 for silencers on average, under a positive-to-control sample ratio of 1:9 (Figure 1C).

Applying the TREDNet model to distal-RE adSNVs, we identified 1,457 silencer and 3,084 enhancer adSNVs in the DLPFC (Table S2). These putative regulatory adSNVs (dubbed radSNVs) formed the primary focus of this study. On average, a radSNV overlaps with 1.5 transcription factor (TF) ChIP-seq peaks detected for the neuronal cell line SK-N-SH from the ENCODE project ^29^. This overlap represents a threefold enrichment compared to non-exon-promoter adSNVs and common SNVs archived in the dbSNP database ^30^ (binomial test *P* < 10^−10^, Figure 1D). Furthermore, radSNVs exhibit a higher density of chromatin contacts detected in DLPFC cells ^31^ compared to both non-exon-promoter adSNVs and common SNVs (*P* < 10^−10^, Figure 1E). Next, 3.1% of radSNVs reside in genomic regions conserved across 100 vertebrate species ^32^, significantly exceeding 1.9% and 2.3% observed for non-exon-promoter adSNVs and common SNVs, respectively, (*P* ≤ 0.005, Figure 1F), which is indicative of the evolutionary significance of their host regulatory elements. radSNVs also display higher GWAS AD association significance levels than other adSNVs (*P* ≤ 0.006, Figure S1). Combined, these results underscore the regulatory and phenotypic importance of radSNVs in brain cells.

Further analysis revealed distinct distribution patterns of silencer and enhancer radSNVs in AD susceptibility loci. We herein identified 12 loci enriched exclusively with silencer radSNVs (referred to as SL loci), 43 loci predominantly with enhancer radSNVs (EN loci), and 12 loci with both enhancer and silencer radSNVs (ENSL loci, Figure 1G and Table S1). Of note, an AD susceptibility locus often spans multiple adjacent AD-associated gene loci. ENSL loci featuring an interplay between activating and repressive regulatory elements encompass 255 AD-associated gene loci, including *ABCA7*, *APOE*, *APOC2*, *BCL3*, *CLU*, *CR1*, *CRHR1, MAPT*, *PTK2B*, etc. ^33^ SL loci comprise 115 gene loci in total, including the genes that have been implicated in neuroinflammation and neuroimmune dysregulation in AD brains, such as *HLA-D*, *MS4A6A*, *TREM2*, *TREM4L*, and *USP6NL* ^33^. EN loci include 188 gene loci, featuring prominent AD-associated genes like *BIN1*, *BCL7*, and *CASS4*.

### AD-associated regulatory SVNs lead to substantial gene upregulation in SL loci

To investigate the biological roles associated with these locus classes, we turned to the genes in these loci. Although proximal genes may not fully account for long-range regulatory targets, they are commonly used to infer the biological functions of regulatory loci ^34^. Notably, genes within SL loci exhibit lower expression in the healthy DLPFC than all assayed genes (Wilcoxon rank-sum test *P* = 7 × 10^−7^), and those in EN loci, ENSL loci or unclassified AD susceptibility loci (represented as UC loci, *P* ≤ 4 × 10^−7^, Figure 2A). In contrast, genes in ENSL loci are expressed at high levels (*P* = 0.04 vs all genes).

**Figure 2.**
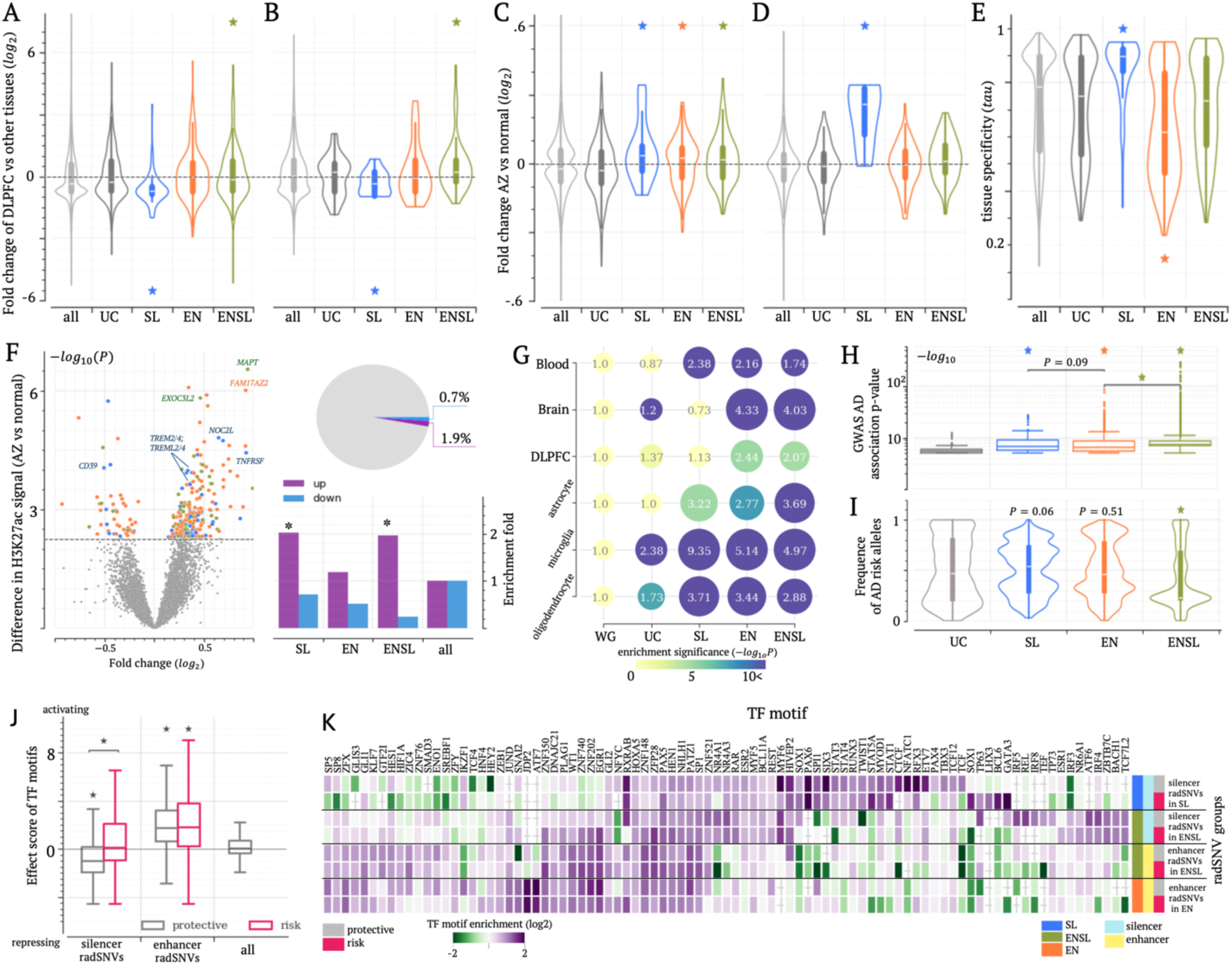
Unique transcriptomic and epigenomic patterns for each locus class. (A, B) Normalized expression levels in the healthy DLPFC for genes associated with different locus classes, linked by (A) genomic proximity and (B) brain eQTL associations. (C, D) Expression fold changes between healthy and AD DLPFCs for genes associated with different classes via (C) proximity and (D) brain eQTLs. (E) Tissue specificity (quantified by *tau*) of genes within different locus classes (see Supplementary Notes). In (A-E), asterisks on the top of violin shapes indicate a significant difference compared to all examined genes. (F) Changes in H3K27ac signals between healthy and AD DLPFCs (the volcano plot in the left panel), and their distribution across different locus classes (the bar plot and the pie chart in the right panel). In the volcano plot, each dot represents a H3K27ac peak. Grey dots indicate a peak with an insignificant change. Blue, orange, and blue dots indicate peaks with significant changes located in SL, EN, and ENSL loci, respectively. Results for H3K27me3 signals are presented in Figure S2. Asterisks on the top of bars indicate a significant difference compared to all H3K27ac peaks. (G) Enrichment of chromatin contacts detected in brain and blood cells, with numbers in dots indicating enrichment levels. Dot sizes and colors denote enrichment level and significance, respectively. (H) Distribution of GWAS association significance (−*log*_10_*P*). The upper and lower whisker edges in these boxplots represent approximately 25% and 75% quartiles of the presented data. (I) Frequency of AD risk alleles among radSNVs. In (H, I), asterisks or p-values on the top of boxes indicate a significant difference compared to UC-locus adSNVs. (J) Effect scores for TF motifs enriched in different radSNV groups with protective and risk alleles, compared to those of all TF motifs. Effect scores represent the differences in TF motif densities between H3K27ac and H3K27me3 ChIP-seq peaks in the DLPFC. Asterisks on the top of boxplots indicate a significant difference compared to all TF motifs. The upper and lower whisker edges in these boxplots represent approximately 25% and 75% quartiles of the presented data. (K) TF motif enrichment in radSNVs with protective and risk alleles. ∗: *P* < 10^−5^.

To explore regulatory effects beyond genomic proximity, we utilized brain-cell gene-locus associations documented in two expression quantitative trait loci (eQTL) datasets: the GTEx project ^35^ and a brain single-cell eQTL database ^36^. By correlating genotypic variations with transcriptomic changes, eQTL analyses reveal regulatory interactions between genomic loci and genes. Among brain eQTL genes, those associated with SL-locus radSNVs are expressed at the lowest levels (*P* ≤ 0.03 vs other brain eQTL genes), whereas ENSL-locus-associated genes exhibit the highest expression levels (*P* ≤ 0.009, Figure 2B). These findings, along with those based on proximal genes (Figure 2A), corroborate the silencing and activating effects of SL and ENSL locus radSNVs, respectively.

We further examined differential gene expression between healthy and AD brains ^37^. Unexpectedly, genes in either SL, EN, or ENSL loci are upregulated more frequently in AD brains than other genes (Wilcoxon rank-sum test *P* < 0.05 vs all tested or UC-locus genes, Figure 2C). Strikingly, 60% of SL-locus differentially expressed genes (DEG) are upregulated in AD brains, surpassing the 43% of all DEGs and 53% of DEGs in all AD susceptibility loci (binomial test *P* < 10^−5^). This suggests that SL-locus genes exhibit the largest upregulation levels, likely due to aberrant silencer activity. In further support of this hypothesis, brain eQTL genes associated with SL-locus radSNVs show the highest upregulation in AD brains (Wilcoxon rank-sum test *p* < 0.0008 vs all brain-eQTL genes, Figure 2D), with 100% of these DEGs being upregulated (*P* < 10^−5^ vs 49% and 56% for EN-locus and ENSL-locus eQTL DEGs, respectively). Furthermore, SL-locus genes show the highest tissue specificity among all tested genes (*P* < 10^−8^), whereas ENSL-locus genes exhibit the lowest (*P* < 10^−5^, Figure 2E). These findings suggest AD-protective roles to silencers in SL loci with radSNVs deactivating silencers and leading to disease-associated overexpression of their target genes.

In AD DLPFC, gene upregulation trends align with the alterations in histone modifications embodied by the gain of H3K27ac activity and loss of H3K27me3 activity, together symptomatic of enhancer gains and silencer losses (see Methods, Table S3). For example, 3.4% of H3K27ac peaks within R to ENSL loci exhibit significant intensity increase, representing a twofold enrichment compared to all H3K27ac peaks (binomial test *P* < 10^−10^). Conversely, only 0.2% of H3K27ac peaks in these regions show decreased intensities, a marked depletion compared to 1.2% of all H3K27ac peaks (*P* < 10^−10^, Figure 2F). Similar trends are observed for SL and EN loci (*P* < 10^−5^ vs all H3K27ac). Furthermore, H3K27me3 peaks near SL and ENSL loci feature decreased intensity in AD DLPFC (*P* < 10^−5^ vs all H3K27me3 peaks, Figure S2). For example, H3K27ac signals significantly rise in AD DLPFC at ENSL loci containing *MAPT* and *EXOC2L3* and the SL loci hosting *TREM2* and *TREML4*. H3K27me3 signals decrease in the SL loci containing *MS4A* and *HLA-D* genes in the AD DLPFC. Combined, these results argue for combinatorial enhancer gain and silencer loss playing an additive effect in boosting AD-associated gene upregulation.

Chromatin organization data further highlight divergent regulatory activities across locus classes. Overall, elevated levels of Hi-C contacts were observed in all three classes—SL, ENSL, and EN loci, — and across different brain cells, often significantly greater than the level of contacts in the genome overall (*P* < 10^−5^ vs the genome-wide average), depicting that radSNVs of these three classes are associated with actively up- and down-regulated genes. ENSL and EN loci enriched for enhancer radSNVs, show particularly dense chromatin contacts detected by H3K27ac HiChIP screens in brain cells ^38^ (binomial test *P* < 10^−10^ vs the entire human genome and UC loci, represented by the second row in Figure 2G), while SL loci, primarily comprising silencer radSNVs, exhibit fewer such chromatin contacts (*P* < 10^−10^, Figure 2G). On the other hand, this trend is reversed among chromatin contacts detected in Hi-C assays not restricted to H3K27ac regions. SL loci, alongside EN and ENSL loci, are enriched with chromatin contacts detected using a Hi-C assay for prefrontal cortices ^39^. Notably, using single-cell Hi-C chromatin interactions detected for prefrontal cortices ^16^, we observed that SL loci exhibit the highest densities of these interactions in microglia and oligodendrocytes, key cell types for neuroimmune regulation (*P* < 10^−7^, Figure 2G). This trend was also observed in blood cells (*P* < 10^−7^ SL loci vs all other loci) ^40^, key modulators of immune systems. These findings reinforce the regulatory importance of SL, ENSL, and EN locus classes for brain cells, and highlight specifically elevated regulatory activity in SL loci involved in neuroimmune regulation.

radSNVs in SL, ENSL, and EN loci also exhibit stronger AD associations than those in UC loci (*P* < 10^−30^, Figure 2H), with those in ENSL and SL loci ranking at the top. radSNVs in ENSL loci have the lowest disease allele frequencies (*P* < 10^−50^ vs other radSNVs, Figure 2I), while those in SL loci exhibit the highest (*P* ≤ 0.06 vs other radSNVs). Collectively, while all locus classes contribute to AD pathogenesis, each class features unique transcriptomic, epigenomic, and genotypic signatures, indicating their distinct roles in this disease. Our locus annotations may offer critical insights into the molecular basis of this complex polygenic disease.

### CTCF and REL repression are among key disruptions by silencer radSNVs

To investigate regulatory circuits associated with each locus class, we analyzed the abundance of transcription factor (TF) binding motifs mapped to radSNVs. The regulatory effect of a TF was quantified by comparing the density of its binding motifs in H3K27ac ChIP-seq peaks (indicative of regulatory activation) versus H3K27me3 ChIP-seq peaks (marking repression). A binding motif enrichment in H3K27ac peaks was recorded as a positive effect score, while the opposite enrichment in H3K27me3 peaks—a negative effect score (see Methods). Consistent with their regulatory functions, silencer radSNVs frequently show high enrichment for TF motifs having negative effect scores (*P* = 10^−7^ vs all TF motifs), whereas enhancer radSNVs are typically abundant in TF motifs with positive scores (*P* = 10^−8^, Figure S3).

TF motif enrichment profiles differ across radSNV classes (Figures 2J and S4). For example, CTCF binding motifs are enriched uniquely among silencer radSNVs in SL loci, whereas REL binding motifs are preferentially overrepresented among silencer radSNVs in ENSL loci. These enrichments are attenuated for AD risk alleles. For example, among silencer radSNVs, TF motifs enriched in risk alleles show elevated effect scores compared to those in protective alleles (*P* = 7 × 10^−6^, Figure 2K). It suggests that substitutions at these radSNVs are associated with motif loss for repressive TFs, such as REL, TF63, SIX3, and CTCF (Figure S5), and/or the gain of activating motifs (for instance, ERG1, GLI1, and SPI1), during AD progression. This trend aligns with the pattern of gene upregulation observed in SL loci (Figures 2C and 2D), likely due to the loss of silencing activity. Together, radSNVs across locus classes exhibit diverse sequence features, with each class recruiting a distinct set of TFs and regulatory networks. These regulatory networks are rewired during AD progression.

### SL-locus radSNVs are strongly associated with immune responses and autoimmune phenotypes

To assess the biological role of radSNVs in different locus classes, we used the Genomic Regions Enrichment of Annotations Tool (GREAT) ^34^. Notably, with minor overlap across classes, each locus class has unique biological functions (Figure 3A). SL loci are uniquely associated with immune-related processes, including immune defense and cellular responses to interferon-gamma (INF*γ*, GREAT *P* < 10^−20^). INF*γ*, a critical pro-inflammatory cytokine for brain defense against latent invaders, has been implicated in microglial hyperactivation in AD brains ^41^. In contrast, EN loci are associated with lipid tube assembly, whereas ENSL loci preferentially govern memory processes and synaptic activity (Figure 3A).

**Figure 3.**
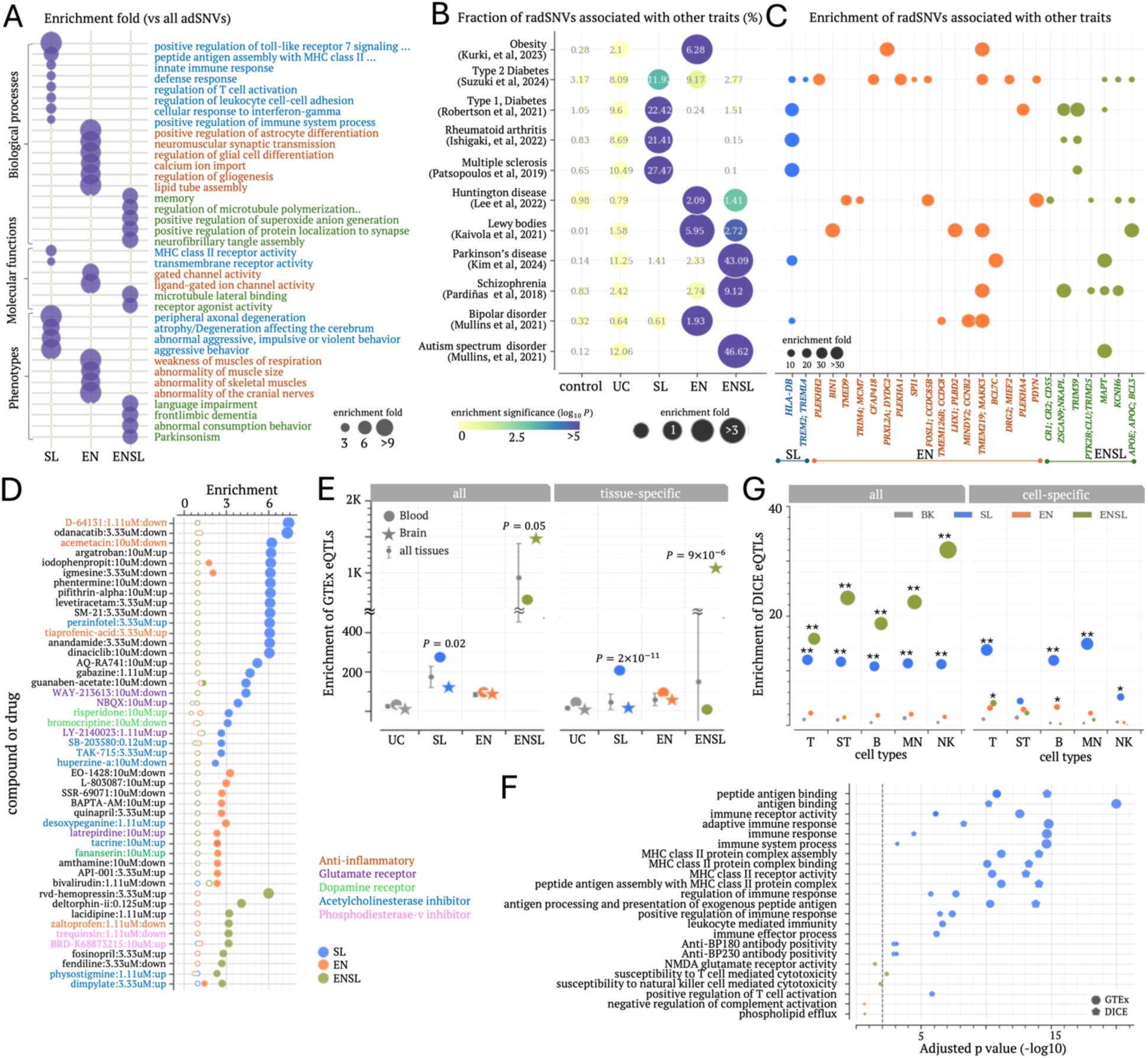
Functional distinctions across locus classes. (A) Functional associations (enrichment folds) of radSNVs across locus classes, based on GREAT analysis. Only significant associations are shown here. (B) Proportions of radSNVs associated with other diseases (%) as indicated by the numbers in dots. Dot colors indicate enrichment levels compared to all GWAS SNVs (represented as WG here). (C) Enrichment of radSNVs associated with other diseases across different loci compared to WG. Dot sizes indicate enrichment folds. Only significant enrichments are shown here. (D) Enrichment of compound-responsive genes in locus classes, with hollow dots indicating insignificant enrichments compared to genome-wide averages. (E) Enrichment of radSNVs across locus classes among GTEx eQTL variants in brain and blood. Silencer and enhancer radSNVs exhibit comparable enrichment levels (Figure S6). Bars are the summarization of enrichments in all tissues. The upper and lower whisker edges in these bars represent approximately 25% and 75% quartiles of these enrichments. P-values indicate the difference significance compared to enrichments in all tissues. The upper and lower whisker edges in these boxplots represent approximately 25% and 75% quartiles of the presented data. (F) Functional analysis of eQTL genes. (G) Enrichment of DICE eQTLs among radSNVs. Asterisks indicate the enrichment significance. ∗∗ : *P* < 10^−5^ and ∗ : *P* < 0.05.

To explore the phenotypic influence of radSNVs in these locus classes, we examined their associations with eleven diseases reportedly relevant to AD. They included six neurological disorders – Parkinson’s disease (PD) ^42^, bipolar disorder ^43^, autism spectrum disorder (ASD), schizophrenia ^44^, Lewy body disease ^45^, and Huntington’s disease ^46^ – as well as three autoimmune diseases (multiple sclerosis ^47^, type 1 diabetes ^48^ and Rheumatoid arthritis ^49^) and two metabolic conditions (type 2 diabetes ^50^ and obesity ^51^). Notably, over 20% of radSNVs in SL loci are associated with all three tested autoimmune diseases, far exceeding those observed among common SNVs or other radSNVs (binomial test *P* < 10^−16^, Figure 3B). These enrichments underscore the pivotal role of SL-locus radSNVs in immune system regulation, aligning with the results from GREAT (Figure 3A). In contrast, ENLS-locus radSNVs are frequently associated with ASD and PD (*P* < 10^−20^ vs GWAS SNVs or other radSNVs, Figure 3B, see Methods), two neurological disorders linked to tau pathology ^52^, which is also supported by the GREAT’s findings. Meanwhile, EN-locus radSNVs are frequently associated with Lewy body and Huntington’s diseases, with over-twofold enrichments compared to other radSNVs (*P* < 10^−5^ vs GWAS SNVs or other radSNVs, Figure 3B). These findings demonstrate distinct phenotypic contributions of each locus class, with SL loci prominently linked to autoimmune disorders, with either altered or hyperactive immune system activity involving corresponding genes being a risk factor for AD.

To further interrogate AD genetic structures shared with other diseases, we extended this analysis to individual loci (see Methods). The SL locus encompassing *HLA-D* genes, which are essential for coordinating immune response, is highly enriched for SNVs associated with PD and tested autoimmune diseases (enrichment folds > 24 vs GWAS SNVs, *P* < 10^−13^, Figure 3C), highlighting the dysregulation of these genes as the molecular basis shared between AD and these diseases. Similarly, the ENSL locus hosting *MAPT* (encoding tau protein) is identified as a genetic link shared by AD, ASD, and PD. The ENSL *APOE* and the EN *BIN1* loci are associated with Lewy body disease, which pinpoints genetic overlaps between these dementia types (Figure 3C). Collectively, this mapping delineates shared genetic underpinnings between AD and other diseases, offering genetic and molecular insights for further investigation.

To explore cellular responses modulated by these locus classes, we utilized gene expression data from the L1000 project ^53^, which catalogs genes significantly up- or down-regulated by thousands of small molecule perturbagens in over 200 cell types, including eight neuronal cell types. Each locus class exhibits unique perturbagen response profiles, with slight overlap among other classes (Figure 3D). SL-locus genes are often responsive to acetylcholinesterase inhibitors (e.g., perzinfotel and huperzine-a) and glutamate receptor antagonists (such as NBQX and LY-2140023, *P* < 10^−7^ vs all tested genes), both compound classes undergoing investigation for AD treatment ^4^. EN-locus genes are enriched among those regulated by latrepirdine and tacrine, both approved for AD treatment. ENSL-locus genes are often modulated by physostigmine (another acetylcholinesterase inhibitor) and phosphodiesterase-v inhibitors (e.g., trequinsin and BRD-K68873215), a drug class with potential for AD treatment pending further validation ^54^. Furthermore, SL-locus genes are frequently downregulated by three anti-inflammatory compounds (D-64131, acemetacin, and tiaprofenic-acid, *P* < 10^−7^ vs all tested genes), with top-ranked enrichment levels (Figure 3D), consistent with experimental anti-inflammatory strategies for mitigating AD risk ^55^.

Overall, all locus classes demonstrate robust biological and pathophysiological associations with AD compared to the whole genome or UC loci, each displaying distinct functional specializations. SL loci are predominantly engaged in immune-related processes; ENSL loci are linked to tau pathology; and EN loci contribute to metabolic regulation. Our results suggest a potential for developing personalized treatment of AD based on the SL/ENSL/EN locus profiles of patients, targeting the specific pathways corresponding to a genetic passport of an individual.

### eQTLs confirm the primary roles of SL-locus radSNVs in regulating immune systems

To investigate the cellular mechanism influenced by radSNVs, we utilized eQTL data, which capture the impact of non-coding SNVs on gene transcription in specific tissues or cell types. A significant overlap with eQTLs suggests an important regulatory role of variants under investigation. Using eQTL data from the GTEx project ^35^, which encompasses eQTLs for 24 distinct tissues (including brain and blood, see Methods), we observed a pronounced co-localization of SL-locus radSNVs with blood eQTLs (Student’s *t*-test *P* = 0.02 vs all tissues). In contrast, ENSL-locus radSNVs preferentially coincide with brain eQTLs (*P* = 0.05, Figure 3E). These enrichment trends persist when analyzing separately for silencer and enhancer radSNVs delineating further the neurological and immune system components of AD into the identified locus classes and regulatory types (Figure S6).

To further refine our understanding of tissue-specific effects of radSNVs, we focused only on eQTLs unique to individual tissues—an analysis that separates general regulatory effects from tissue-specific regulatory interactions. This analysis reveals a further heightened enrichment of SL-locus radSNVs in blood-specific eQTLs and ENSL-locus radSNVs in brain-specific eQTLs (*P* = 2 × 10^−11^ and *P* = 9 × 10^−6^ vs all tissues for SL and ENSL loci, respectively, Figure 3E). Interestingly, present yet not necessarily overly abundant brain eQTL genes associated with SL-locus radSNVs are predominantly associated with immune-related processes, such as immune response and immune receptor activity (adjusted *P* < 10^−10^, Figure 3F), nevertheless, as assessed using the g:Profiler ^56^. These genes often respond to anti-BP180 antibodies (g:Profiler adjusted *P* = 0.00004), a class of antibodies correlated with the incidence and severity of dementia ^57^. These findings, in line with analyses using the GREAT tool and GWAS associations (Figures 3A and B), reinforce the regulatory significance of SL-locus radSNVs in blood cells and ENSL-locus radSNVs in brain cells.

To further elucidate the regulatory impact of radSNVs on immune system genes and the corresponding cellular specificity, we leveraged the data from the database of immune cell eQTLs (DICE, Figure S7) ^58^. Both ENSL- and SL-locus radSNVs show substantial enrichment for DICE eQTLs across blood cell types, including T, stimulated T, B, monocyte, and natural killer cells, with over 10-fold enrichments compared to all GWAS SNVs or other radSNVs (*P* < 2 × 10^−11^, Figure 3G). Interestingly, SL-locus radSNVs (but not ENSL-locus and EN-locus radSNVs) are uniquely enriched in eQTLs specific to a single cell type, presenting a cell-type specific and centric impact of these variants. For example, these radSNVs coincide with monocyte-specific eQTLs three times more often than common SNVs or other radSNVs (*P* < 10^−17^, Figure 3G). Moreover, DICE eQTL genes associated with SL-locus radSNVs, such as those in the *HLA-D* family, are enriched in immune regulation pathways (g:Profiler adjusted *P* < 4 × 10^−4^, Figure 3F). These findings, mirroring those from GTEx-based and functional annotation analyses (Figures 3A), further highlight SL-locus radSNVs as key modulators of neuroimmune systems with granular cellular specificity.

### SL-locus radSNVs are selectively linked to gene upregulation in AD microglia

Microglia, the brain’s resident immune cells derived from monocytes, are key players in responding to harmful stimuli ^59^. The strong association of SL-locus genes with immune and hematopoietic systems, particularly monocytes, prompted an investigation into their roles in modulating microglial states and functions in healthy and AD DLPFCs. With single-cell transcriptomic data for DLPFCs from the ssREDA data resource ^60^ and a study by S. Morabito et al. ^27^, we clustered differentially expressed genes (DEGs) between healthy and AD cases into eight distinct groups (Figure 4A). Seven of these clusters exhibit cell-specific upregulation patterns in the AD DLPFC. For example, *HLA-DRA/Q*, *EED*, *APOC*, and *TMEM529* are upregulated exclusively in AD microglia, whereas *PTK2B* and *CRHR1* show a strong upregulation level primarily in AD excitatory neurons. The exception cluster (the “pink” in Figure 4A) contains the genes upregulated across multiple cell types in the AD DLPFC, with an example of *MAPT* that is upregulated in AD excitatory and inhibitory neurons, microglia, and oligodendrocytes.

**Figure 4.**
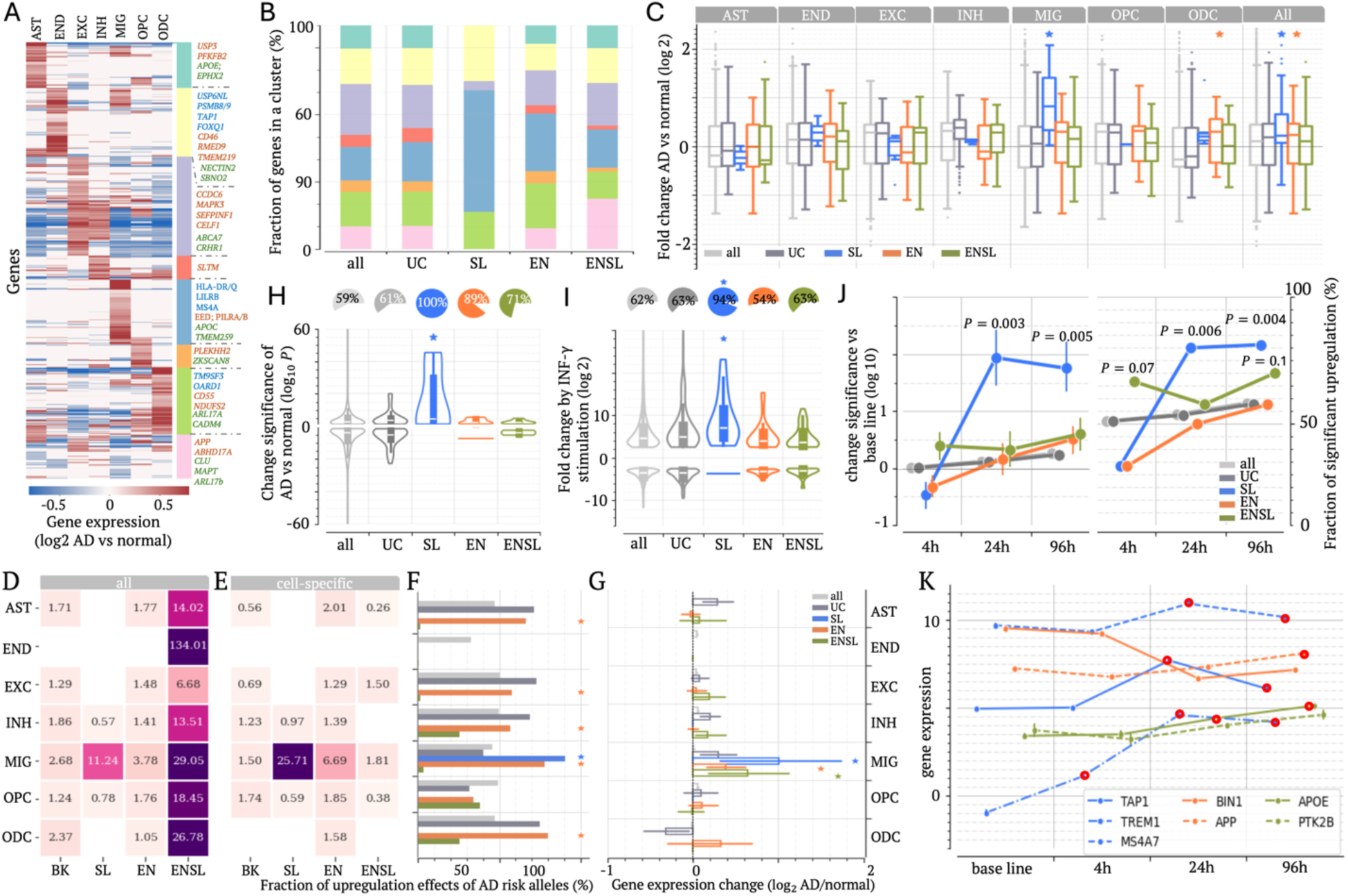
SL-locus radSNVs are associated with gene upregulation in AD microglia. (A) Heatmap illustrating gene clustering based on expression changes in AD across cell types: astrocytes (AST), endothelial cells (END), excitatory neurons (EXC), inhibitory neurons (INH), microglia (MIC), oligodendrocyte progenitor cells (OPC), oligodendrocytes (ODC). (B) Distribution of these expression clusters across locus classes. (C) Gene expression across cell types for different locus classes, with asterisks denoting significant differences compared to all tested genes. Asterisks indicate a significant difference compared to all examined genes. The upper and lower whisker edges in these boxplots represent approximately 25% and 75% quartiles of the presented data. (D, E) Enrichment of brain eQTLs among radSNVs across locus classes, with (D) for all eQTLs and (E) for cell-specific eQTLs. (F) Proportions of eQTL radSNVs where AD risk alleles are associated with increased gene expression. (G) Differential expression of microglia-eQTL genes associated with radSNVs across locus classes in AD microglia. (H) Gene expression differences between healthy and AD microglia (quantified by differential significance −*log*_10_*P*) across locus classes. Positive/negative values represent up-/down-regulations in AD microglia. The pie charts on the top display the fractions of upregulated genes. (I) Gene expression after INF-*γ* stimulation across locus classes, with the pie charts above summarizing the fractions of upregulated genes. (J) Differential gene expression after the stimulation of pre-formed Aβ fibrils. All examined genes (represented by the light grey) are the control for significance analysis. Only significant p-values are shown. The upper and lower whisker edges in error bars represent approximately 25% and 75% quartiles of the presented data. (K) Temporal gene expression profiles after the stimulation of pre-formed Aβ fibrils, with red-circled dots highlighting the significant upregulations compared to untreated baseline levels. In (C, F, G, H, I), asterisks and p-values indicate difference significance compared to all examined genes. ∗ : *P* < 10^−5^.

Of SL-locus genes, 54% are upregulated predominantly in microglia (the blue cluster in Figure 4A), a proportion markedly surpassing the less-than-25% observed for all DEGs or DEGs from other locus classes (binomial test *P* < 10^−7^, Figure 4B). Furthermore, microglia-upregulated DEGs in SL loci show the highest upregulation levels (*P* = 2 × 10^−6^ vs all microglia-upregulated DEGs, Figure 4C). To further validate this trend, we analyzed gene expression profiles in healthy and AD microglia published by N. Sun et al. ^61^, identifying microglia-DEGs. Strikingly, 100% of microglia-DEGs in SL loci are upregulated in AD, significantly exceeding 59% of the genome-wide average and 61-89% seen in other AD susceptibility loci (*P* < 10^−10^, Figure 4D). Similarly, SL-locus upregulated microglia-DEGs exhibit the highest upregulation levels (Student’s *t*-test *P* = 6 × 10^−5^ vs other microglia-upregulated genes, Figure 4D).

To further evaluate the microglia specificity of SL-locus radSNVs, we utilized eQTLs detected across multiple brain cell types, including astrocytes, endothelial cells, excitatory and inhibitory neurons, microglia, oligodendrocytes, and oligodendrocyte progenitor cells ^36^. In contrast to EN- or ENSL-locus radSNVs, SL-locus radSNVs frequently co-localize with microglia eQTLs (binomial test *P* = 2 × 10^−15^ vs common SNVs, Figure 4E), especially those exclusive to microglia (*P* = 10^−22^, Figure 4F). Notably, 70% of SL-locus radSNV eQTLs are microglia-specific (*P* = 0.001 vs 39% of all eQTLs, Figure S8). These findings highlight the unique microglia specificity of SL-locus radSNVs, contrasting ubiquitous roles of ENSL-locus radSNVs across brain cell types. To assess the transcriptional impact of radSNVs in AD, we analyzed eQTL effect sizes, aligning them such that positive values denote gene upregulation in AD brains. Notably, 90.4% of microglia eQTLs co-localizing with SL-locus radSNVs hold positive effect sizes, far exceeding 45.7% observed among microglia eQTLs (*P* < 10^−20^, Figure 4G). Furthermore, microglia-eQTL genes associated with SL-locus radSNVs show the highest upregulation levels in AD microglia (Wilcoxon rank-sum test *P* = 0.001 vs all microglia-eQTL genes, Figure 4H).

Collectively, these findings, corroborated by both single-cell and bulk transcriptomic data (Figures 4B–D) and by both proximal and eQTL genes, underscore the pivotal role of SL-locus genes in driving microglial dysregulation in AD and suggest that aberrant silencing of the corresponding genes in microglia is one of the critical and prominent components of AD, with microglial gene upregulation being emblematic of AD.

### SL-locus genes are predominantly upregulated during microglia inflammation

To further probe how microglia responds to inflammatory stimuli, we analyzed transcriptomic profiles in microglia-like cells generated from induced pluripotent stem cells (iMGLs), both in their basal state and following stimulation with pre-formed A*β* fibrils ^61^ or pro-inflammatory factor INF-γ ^62^. Consistent with their significant upregulation trend in AD microglia (Figures 4C and 4D), SL-locus genes are robustly induced by INF-γ stimulation ^62^. In detail, 94% of SL-locus INF-γ DEGs are upregulated, significantly exceeding the 64% observed in all DEGs (including those in other AD susceptibility loci, *P* < 10^−10^, Figure 4I). Also, INF-γ-upregulated DEGs in SL loci exhibit the highest upregulation levels (Student’s *t*-test *p* = 0.02 vs all INF-γ-upregulated genes, Figure 4I). Notably, 67% of SL-locus INF-γ-upregulated DEGs are also upregulated by pre-formed Aβ fibrils, representing a notable enrichment compared to 27% observed among all INF-γ-upregulated DEGs (*P* = 0.001, Figure S9). These findings highlight the leading role of SL-locus genes in orchestrating microglial responses against diverse inflammatory stimuli.

Analyzing time-resolved transcriptomics of iMGLs exposed to pre-formed Aβ fibrils over the course of 4-96 hours ^61^, we further revealed distinct temporal patterns for different locus classes. SL-locus genes display pronounced upregulation from 24 hours onward, contrasting with the modest early upregulation of ENLS-locus genes at four hours after Aβ seeding and negligible changes of EN-locus genes throughout the course (the latter one aligns with insignificant changes between healthy and AD brains of EN-locus genes, as presented in Figure 4C). Specifically, 65% of ENSL-locus DEGs are upregulated at 4 hours after the pre-formed Aβ exposure, whereas over 80% of SL-locus DEGs are upregulated from 24 hours onward, both exceeding the expectations among all DEGs (*P* = 0.07 and *P* = 0.006, respectively, Figure 4J). For example, SL-locus genes *TAP1*, *TREM1*, and *MS4A7* are upregulated from 4 hours and beyond (Student’s *t*-test, *P* < 10^−4^), whereas the EN-locus gene *BIN1* is downregulated throughout this course (Figure 4K). Together, different locus classes exhibit diverse cell- and time-specific response patterns during AD progression, with SL-locus genes sustaining robust upregulation in both AD and pro-inflammatory-stimulated microglia.

### SL-locus genes show elevated expression levels in A*β*-predominant AD subtypes C1 and C2

The profound genetic and clinical heterogeneity of late-onset AD has driven its stratification into molecularly defined subtypes. Transcriptome analyses of hundreds of human brains have delineated five AD subtypes (A, B1, B2, C1, and C2), each exhibiting unique molecular signatures while sharing similarities in disease severity, biological sex, and the age of onset and death ^9^. Subtypes A, B1, and B2 are marked by tau protein dysregulation, with subtype A uniquely demonstrating the resilience to neurofibrillary tangles. Subtypes C1 and C2 are distinguished by the overrepresentation of A*β* binding and aggregation.

Given the functional diversity of locus classes, we examined their correspondence with AD subtypes. Genes in SL loci are overexpressed in subtype C (including C1 and C2, Wilcoxon rank sum test *P* < 0.0009 vs all genes, Figure 5A). Subtypes B1 and B2 exhibit increased expression of genes in ENSL loci (*P* < 0.0009), whereas subtype A is characterized by low expression of genes across all locus classes (*P* < 0.03, Figure 5A). These expression patterns are reflected among genes associated with radSNVs via brain eQTLs archived in the GTEx ^35^ or the brain-cell eQTL databases ^36^. For instance, eQTL genes associated with SL-locus radSNVs are overexpressed in subtypes C1 and C2 (*P* < 0.003), whereas those with ENSL loci are upregulated in subtypes B1 and B2 (*P* < 10^−5^). Subtype A shows downregulation of all these genes (*P* < 0.006, Figure 5B). For example, the genes *USP6NL* and *HLA-DQ* family, associated with SL loci by either proximity or eQTLs, exhibit elevated expression levels in subtypes C1 and C2 (Student’s *t*-test *P* < 0.006 vs other subtypes). *MAPT*, an ENSL-locus gene via proximity or eQTLs, is overexpressed in subtypes B1 and B2 (*P* = 0.04, Figure 5C).

**Figure 5.**
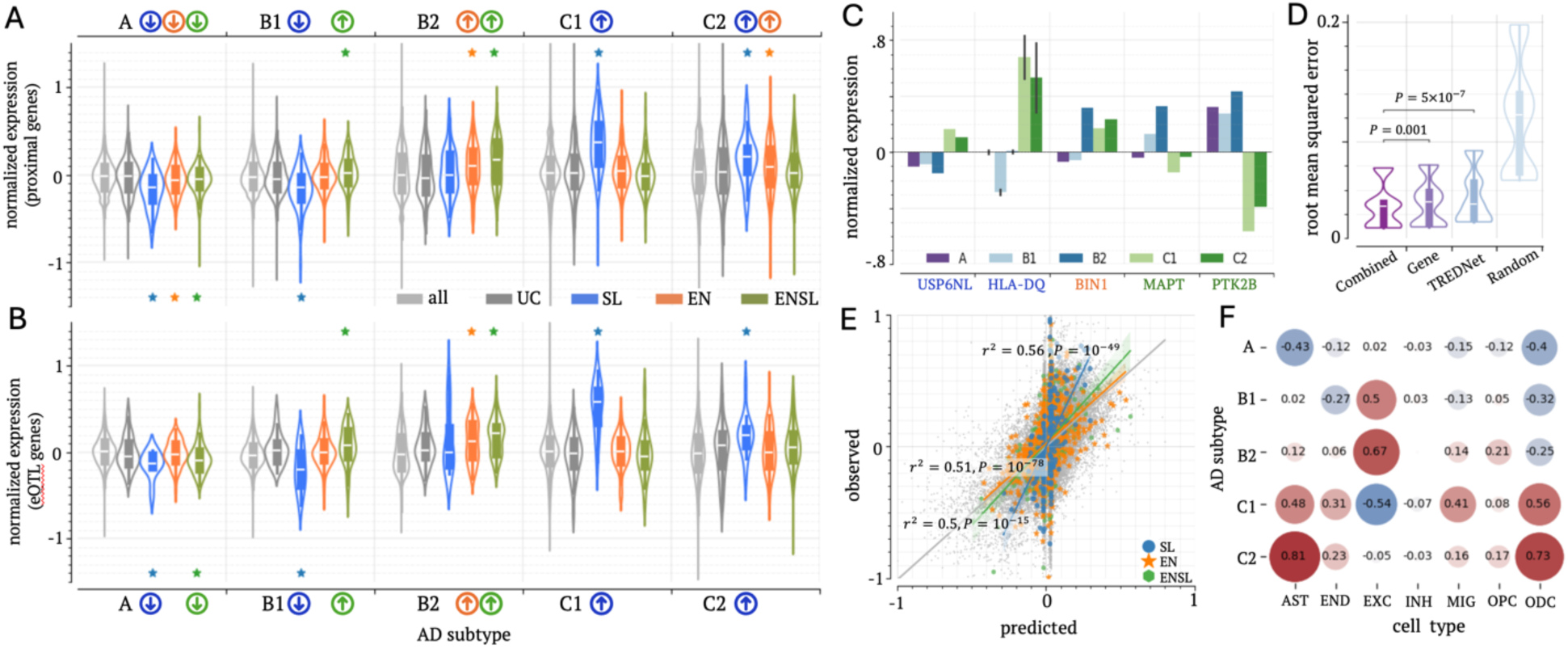
Each locus class shows a unique subtype specificity. (A, B) Subtype-specific gene expression associated with locus classes by (A) proximity and (B) brain eQTL associations. Asterisks indicate significant differences compared to all tested genes. ∗ : *P* < 10^−5^. (C) Subtype-specific expression levels of example genes. In the case of HLA-DQ gene family, the upper and lower whisker edges in these bars represent approximately 25% and 75% quartiles of the presented data. (D) RMSE values calculated over test samples for linear regression models in independent 100 trials. (E) Comparison of observed and predicted gene expression across locus classes. (F) Average weights in linear regression models built on cell-specific gene expression data and enhancer/silencer profiles. Cell types include astrocytes (AST), endothelial cells (END), excitatory neurons (EXC), inhibitory neurons (INH), microglia (MIC), oligodendrocyte progenitor cells (OPC), and oligodendrocytes (ODC). Weights in other models are summarized in Figure S11.

To gain further insights into the cellular characteristics of AD subtypes, we trained lasso regression models to predict gene expression levels for each AD subtype using cell-specific enhancer/silencer activity predictions across seven major brain cell types: astrocytes, endothelial cells, excitatory and inhibitory neurons, microglia, oligodendrocytes, and oligodendrocyte progenitor cells (see Methods). Across 100 independent trials, our models achieved an average root mean square error (RMSE) of 0.051, largely outperforming random shuffling (average RMSE = 0.12, Student’s *t*-test *P* < 10^−50^, Figure 5D) and performing comparably to models built on DLPFC cell-type gene expression data (average RMSE = 0.050, *P* = 0.001). Furthermore, combining enhancer/silencer with cell-specific gene expression further improved regression performance (average RMSE=0.047, *P* ≤ 0.002 vs other models). High regression accuracies were sustained across AD subtypes (Figure S10) and locus classes, with predicted expression correlating with observed expression at 0.56 for SL-locus genes (*P* = 10^−49^, Figure 5E). Collectively, these results underscore that our enhancer/silencer profiles encompass essential regulatory components across cell types in the DLPFC, capturing the gene regulation patterns underlying different biological pathways and AD subtypes.

Furthermore, weights from the regression models were used to establish the cellular activity profiles for AD subtypes. For example, subtypes C1 and C2 correlate positively with the activity of immune-related cells, including astrocytes, microglia, oligodendrocytes, and endothelial cells, but negatively with the activity of excitatory neurons (Figure 5F). These findings align with reports of increased microglia and astrocyte populations as well as substantial neuron loss in patients of these subtypes ^9^. They are commonly diagnosed with hyperactive astrocytosis and microgliosis in response to the excessive aggregation of A*β*. In addition, subtypes B1 and B2 show heightened excitatory neuron activity alongside moderate oligodendrocyte loss, whereas subtype A exhibits negative correlations with astrocyte and oligodendrocyte activity (Figure 5F), supporting a distinct cellular activity profile for each AD subtype. Notably, the models built on different data types (i.e., enhancer/silencer profiles, cell-specific gene expressions, and the combination of them) show similar weights (Figures 5F and S11), validating the robustness of the TREDNet in predicting cell-specific enhancer/silencer activities, as it translates to the separation of AD subtypes.

### Deep learning identifies AD causal variants by accurately quantifying their regulatory impacts

Beyond annotating the function of radSNVs, the TREDNet model was applied to assess the regulatory impact due to variants. Building on our prior study ^63^, these impacts are quantified as the difference in TREDNet-derived prediction scores between protective and risk alleles, denoted as Δactivity (see Methods). A positive Δactivity indicates that the variant increases activation or reduces repression strength, whereas a negative value reflects the opposite effect. radSNVs with significant Δactivity are considered causal for AD ^63^.

Identified putative causal radSNVs overlap with transcription factor (TF) ChIP-seq peaks in the neuronal cell line SK-N-SH more frequently than common SNVs, adSNVs, or other radSNVs (binomial test *P* < 10^−10^), second only to promoter adSNVs (Figure 6A). Similarly, these putatively causal radSNVs are enriched in TF binding motifs significantly enriched in H3K27ac or H3K27me3 ChIP-seq peaks in the DLPFC (*P* < 10^−10^, Figure 6A, see Methods). Furthermore, 5.5% of identified putatively causal radSNVs reside in the regions conserved across 100 vertebrate species ^32^, significantly surpassing that for common SNVs (2.8%) or radSNVs overall (3.1%, *P* = 3 × 10^−5^, Figure 6B). This conservation level trails only that of SNVs in exon and promoter regions, both of which are known for high evolutionary conservation due to their functional role in cellular biology.

**Figure 6.**
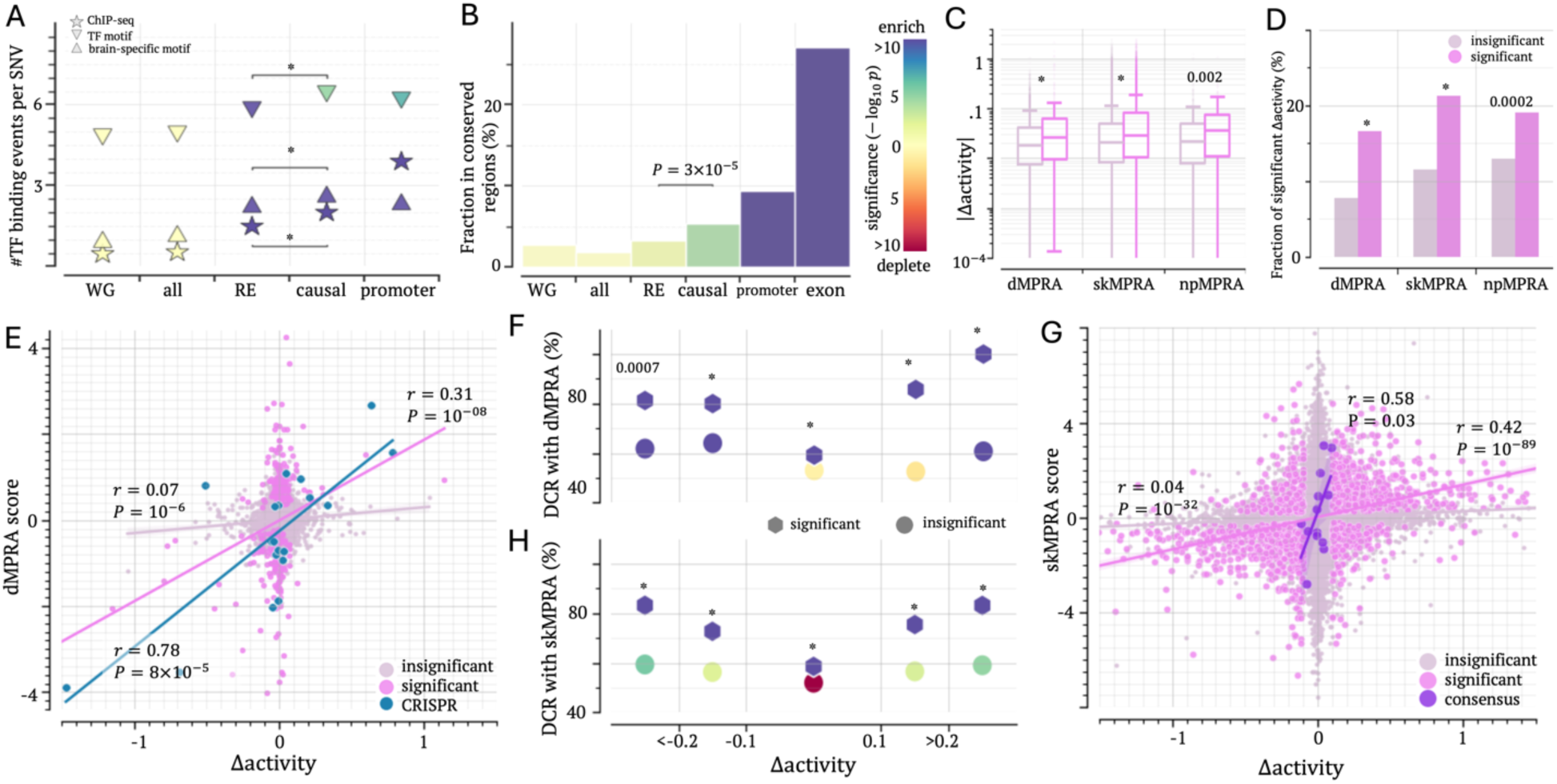
Accurate prediction of regulatory influence of SNVs. (A) Density of TF binding events across variant groups. Asterisks indicate significant enrichments compared to whole-genome common SNVs (WG). (B) Proportions of variants located in evolutionarily conserved regions, with marker and bar colors reflecting statistical significance compared to WG. Colors of markers and bars indicate the degree of difference compared to common SNVs. Asterisks indicate significant differences. (C) Distribution of |Δactivity| values, and (D) fraction of significant Δactivity scores for the variants having significant and insignificant MPRA scores. The upper and lower whisker edges in these boxplots represent approximately 25% and 75% quartiles of the presented data. Asterisks indicate significant differences between significant and insignificant MPRA SNVs. (E) Correlation between Δactivity and dMPRA scores. “CRISPR” represents the variants validated in CRISPR experiments. (F) DCRs between dMPRA and Δactivity scores for variants, stratified by Δactivity. (G) Correlation between DLPFC Δactivity and skMPRA scores. (H) DCRs between skMPRA and Δactivity scores for variants, stratified by Δactivity. The correlation between SK-N-SH Δactivity and skMPRA scores is presented in Figure S12. In F and H, asterisks represent significant differences between significant and insignificant MPRA SNVs. In A, C, F, and H, ∗ : *p* < 10^−5^.

To directly evaluate the ability of Δactivity scores to predict changes with phenotypic impact, we utilized MPRA results which assess the transcriptional activity of assayed DNA sequences. Differences in MPRA outcomes between alleles measure the alteration in regulatory strength due to variants. We utilized MPRA data for thousands of dementia-associated SNVs (dMPRA SNVs), which correlate strongly with CRISPR-based validations in stem-cell-derived brain cells despite being conducted in neuroepithelial-like human embryonic kidney 293T cells (HEK293T) ^19^. Significant dMPRA SNVs exhibit higher absolute Δactivity (|Δactivity|) than insignificant dMPRA SNVs (Wilcoxon rank-sum *P* = 10^−5^, Figure 6C). Of the significant dMPRA SNVs, 16.5% have significant Δactivity scores, representing a twofold enrichment compared to the 7.7% observed among insignificant dMPRA SNVs (binomial test *P* = 10^−76^, Figure 6D).

Critically, Δactivity scores significantly positively correlate with dMPRA outcomes. While the correlation coefficient is modest for insignificant dMPRA SNVs (*r* = 0.07, *P* = 10^−6^, Figure 6E), it increases substantially to *r* = 0.31 (*P* = 10^−8^) for significant dMPRA SNVs, and rises further to *r* = 0.78 (*P* = 8 × 10^−5^) for CRISPR-validated significant dMPRA SNVs. Overall, 63.4% of dMPRA SNVs show directional concordance between Δactivity and dMPRA scores. This directional-concordance rate (namely, DCR) escalates with increasing |Δactivity|, peaking at 88% for |Δactivity| > 0.2 (Figure 6F). Among 10 CRISPR-validated radSNVs, the DCR reaches 90% (Table S4), consistently affirming the robustness of Δactivity scores, which thus can be used to judge the regulatory impact of sequence variants, especially at large absolute value Δactivity scores.

We further validated Δactivity using MPRA data from the neuronal cell line SK-N-SH (termed skMPRA here) published by Alan et al ^20^. Similar to dMPRA SNVs, skMPRA SNV outcomes significantly positively correlate with Δactivity scores. Of the significant skMPRA SNVs, 21% have significant Δactivity scores, representing a twofold enrichment compared to insignificant SNVs (binomial *P* = 10^−164^, Figure 6D). Correlation coefficients between Δactivity and skMPRA SNV scores increase from *r* = 0.04 (*P* = 10^−32^) for insignificant skMPRA SNVs to *r* = 0.42 (*P* = 10^−89^) for significant ones (Figure 6G). The DCR with skMPRA SNV scores approaches 74.3% for significant Δactivity scores (*P* = 10^−110^ vs 52.7% of insignificant Δactivity) and further rises to 83.2% for those with |Δactivity| > 0.2 (Figure 6H). These patterns closely mirror those observed with dMPRA outcomes, reinforcing the reliability of Δactivity scores for evaluating the impact of brain variants.

Of 181 SNVs measured in both dMPRA and skMPRA, 93 SNVs (51%) displayed directional concordance, denoted as consensus SNVs. For them, Δactivity scores correlate significantly with skMPRA measurements (*r* = 0.58, *P* = 0.03), with a DCR of 69% (Figure 6G). The TREDNet model built for SK-H-SH cells in our previous study ^63^ marginally outperforms the DLPFC model (albeit not significantly, Figure S12), demonstrating the ability of TREDNet models to capture cell-specific regulatory dynamics, albeit to a limited extent.

Further validations using MPRA data from human neural progenitors (represented by npMPRA) published by McAfee et al.^21^ also corroborate these findings. A significant correlation of *r* = 0.54 (*P* = 0.005) and a DCR of 72.2% among significant npMPRA SNVs (Figure S13) underscore the robustness of Δactivity scores. Together, validations across three independent brain-related MPRA studies consistently demonstrate the reliability of Δactivity scores in capturing the regulatory effect of SNVs in brain cells, establishing Δactivity scores as a robust metric for prioritizing causal radSNVs for AD.

### Δ*activity* identifies rs636317, disrupting a CTCF binding event, as an AD causal variant in a SL locus

The membrane-spanning 4-domains (*MS4A*) gene cluster, for example, *MS4A6A* and *MS4A4A*, is a key modulator for immune cell activities in the brain ^64^. This genomic locus, harboring numerous adSNVs ^65^, is enriched with DLPFC silencers and is therefore categorized as an SL locus (Figure 7A). This annotation aligns with the low expression of these genes in the healthy human brain ^66^.

**Figure 7.**
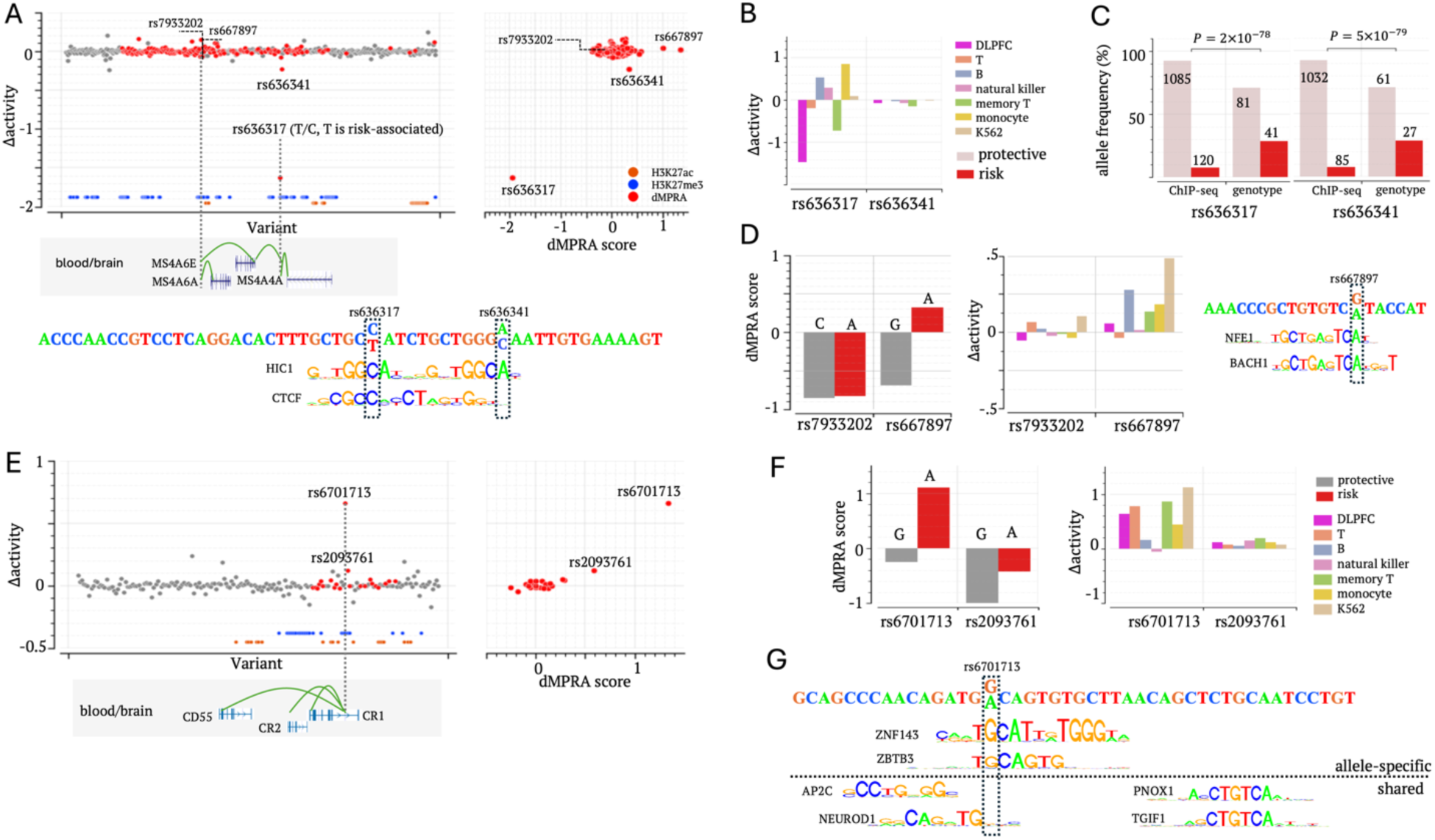
Prioritizing radSNVs in the *MS4A* and *CR1* loci. (A) Δactivity and dMPRA scores in the *MS4A* locus. Left: Δactivity scores; right: comparison of Δactivity (y-axis) with dMPRA (x-axis). (B) Δactivity scores for rs636317 and rs636341 in the DLPFC and blood cells. (C) Allele frequencies for rs636317 and rs636341 in CTCF ChIP-seq reads and the study cohort. The numbers in/above bars are occurrence of alleles. (D) dMPRA results (the left panel) and Δactivity scores (the right panel) for rs7933202 and rs667897. (E) Δactivity and dMPRA scores in the *CR1* locus. Left: Δactivity scores; right: comparison of Δactivity (y-axis) with dMPRA (x-axis). (F) dMPRA outcomes (the left panel) and Δactivity scores (the right panel) for rs6701713 and rs2093761. (G) Δactivity and dMPRA scores in the *CR1* locus. (F) TF motif mapping analysis for rs6701713.

Among the radSNVs in this locus, rs636317 exhibits the highest |Δ*activity*|, perfectly reflecting its leading dMPRA score among 168 dMPRA SNVs in this region (Figure 7A). Rs636317 has been marked as a likely AD causal SNV, with the allele C acting as protective and the allele T conferring AD risk. The allele C is significantly overrepresented in CTCF ChIP-seq reads (90% of reads carrying C vs its allele frequency of 67% in brain samples, *P* = 2 × 10^−78^, Figures 7B). This result aligns with the finding that the allele T disrupts a CTCF binding site ^28,65^ (Figure 7A). Despite these converging findings, the regulatory impact of the substitution at rs636317 remains unclear, as both overexpression and underexpression of *MS4A* genes have been hinted to exacerbate AD progression ^67^. Moreover, while the risk allele T is associated with reduced transcriptional activation in microglia and brain cells (as measured in dMPRA experiments), it corresponds to the increased expression of *MS4A6A* in monocytes ^65^ and the GTEx blood cells ^35^. This apparent discrepancy may reflect the pleiotropic nature of rs636317 and the context-dependent function of its host element. To probe this further, we used TREDNet models to predict the regulatory effect of rs636317 in the DLPFC and blood cell types (such as T and B cells, monocytes, natural killer cells, etc.). We had previously demonstrated that Δ*activity* scores significantly correlate with MPRA outcomes in blood cell types, reliably prioritizing regulatory SNVs for autoimmune diseases such as type 1 diabetes ^63^. Here, rs636317 has a significantly negative Δ*activity* in the DLPFC but positive Δ*activity* scores in B cells and monocytes (Figure 7C). These predictions are consistent with with the dMPRA measurements (Figure 7A) and eQTL associations in GTEx blood cells and macrophage ^65^, confirming the reliability of Δ*activity* scores, allowing us to conclude a pleotropic influence of this variant in DLPFC and immune system cells.

Interestingly, rs636341, a nearby radSNV located 11 bp from rs636317, shows negligible Δactivity in both the DLPFC (consistent with its insignificant dMPRA score) and blood cells (Figure 7B). Due to their genetic and genomic proximity, rs636317 and rs636341 show similar AD GWAS association levels and allele bias in CTCF ChIP-seq reads (Figure 7C). This highlights the inherent difficulty in pinpointing causal SNVs amidst nearby variants and thus underscores the effectiveness of TREDNet models (broadly, deep learning models) and MPRA assays for accurately identifying causative SNVs. In addition, AD risk alleles of rs636317 and rs636341 disrupt the binding motif of HIC1 (Figure 7A), a repressor and regulator of chromosomal stability ^68^. These disruptions potentially compromise local chromatin architecture, conferring the risk of *MS4A* dysregulation, in AD patients. While further validation is necessary, this hypothesis offers an alternative therapeutic target for AD in the *MS4A* locus.

Finally, in the *MS4A* locus, a similar trend emerges for rs667897 and rs7933202, located 53 bps apart. The risk allele at rs667897 introduces a binding site for NRE, potentially thereby augmenting *MS4A6A* expression, whereas the substitution at rs7933202 exerts an insignificant effect ^69^. Furthermore, rs667897, rather than rs7933202, has a significant dMPRA score (Figure 7D). All these experimental assessments are successfully captured by Δactivity scores, with rs667897 holding significant Δactivity in blood cell types (such as B cell, monocytes, and K562) but the Δactivity of rs7933202 remaining insignificant across all tested blood cell types and the DLPFC (Figure 7D).

### Candidate AD causal silencer variants in the *CR1* and *USP6NL* loci

Complement receptor 1 (*CR1* or *CD35*), a pivotal component in the innate immune system, is expressed on the surface of blood cells and facilitates the phagocytosis of immune complexes, including A*β*. The *CR1* locus, categorized as an ENSL locus, is enriched with both enhancer and silencer radSNVs. Among these variants, rs6701713 exhibits the largest |Δ*activity*|, consistent with its leading dMPRA score among 22 dMPRA variants (Figures 7E and 7F). The 60bp-long sequence surrounding rs6701713 contains binding motifs for transcriptional repressors, including PNOX1, TGIF1, and NEUROD1 (Figure 7G). The protective allele G at rs6701713 forms binding motifs for ZNF143, a CTCF cofactor, and ZBTB3, a chromatin looping organizer and transcription repressor ^70^. The risk allele A, disrupting these motifs, is associated with increased *CR1* expression across multiple brain regions, such as the frontal cortex and hippocampus (Figure S14). These findings support the rs6701713-hosting element as a silencer, maintaining low expression of *CR1* in the healthy brain. Disruption of this silencer by the G-to-A substitution at rs6701713 is associated with *CR1* upregulation in AD ^71^. All reports support a significant positive Δ*activity* at rs6701713. Our motif analysis further suggests that this regulatory alteration is probably due to the loss of ZNF143 binding.

In the *USP6NL* locus, another SL locus, seven SNVs were probed in dMPRA experiments. Of them, rs7920721 corresponds to the most significant dMPRA score and Δactivity (Figure S15). The risk allele G at rs7920721 weakens the binding motif for NEUROG1, a TF essential for brain development. Additionally, rs12359970, a variant not examined in the dMPRA, exhibits the highest Δactivity in this locus. The AD risk allele G at this variant disrupts a binding motif of HMGA2 – a TF involved in neuron development and AD pathogenesis ^72^, suggesting the potential contribution of this variant to the dysregulation of *USP6NL* in AD.

### Complexity of AD is reflected in combined impact of silencer and enhancer causal variants

PTK2B, a non-receptor tyrosine kinase, is one of the few validated genes for late-onset AD, with diverse roles in neuroinflammation, neuronal development, and synaptic plasticity ^73^. PTK2B downregulation is linked with tau hyperphosphorylation, whereas its overexpression is associated with A*β*-induced phenotypes such as memory impairment and synapse loss ^73,74^. The *PTK2B-CLU* locus, enriched with enhancer and silencer radSNVs, is categorized as an ENSL locus.

Of 18 probed radSNVs, dMPRA experiments identified three significant variants. Two of them – rs755951 and rs1532277 – reach significant Δactivity scores. Rs755951 displays the highest Δactivity and dMPRA scores (Figure 8A). Its risk allele C, corresponding to increased *PTK2B* expression in blood cells (Figure S14), disrupts, at least attenuates, the binding motif of HIC1, a transcriptional repressor and chromatin organizer (Figure 8B). Another significant dMPRA SNV is rs1532277, where the C-to-T substitution introduces binding motifs for TFEC and TFEB (Figure 8C), transcriptional activators associated with neurodegenerative disorders, including AD ^75^.

**Figure 8.**
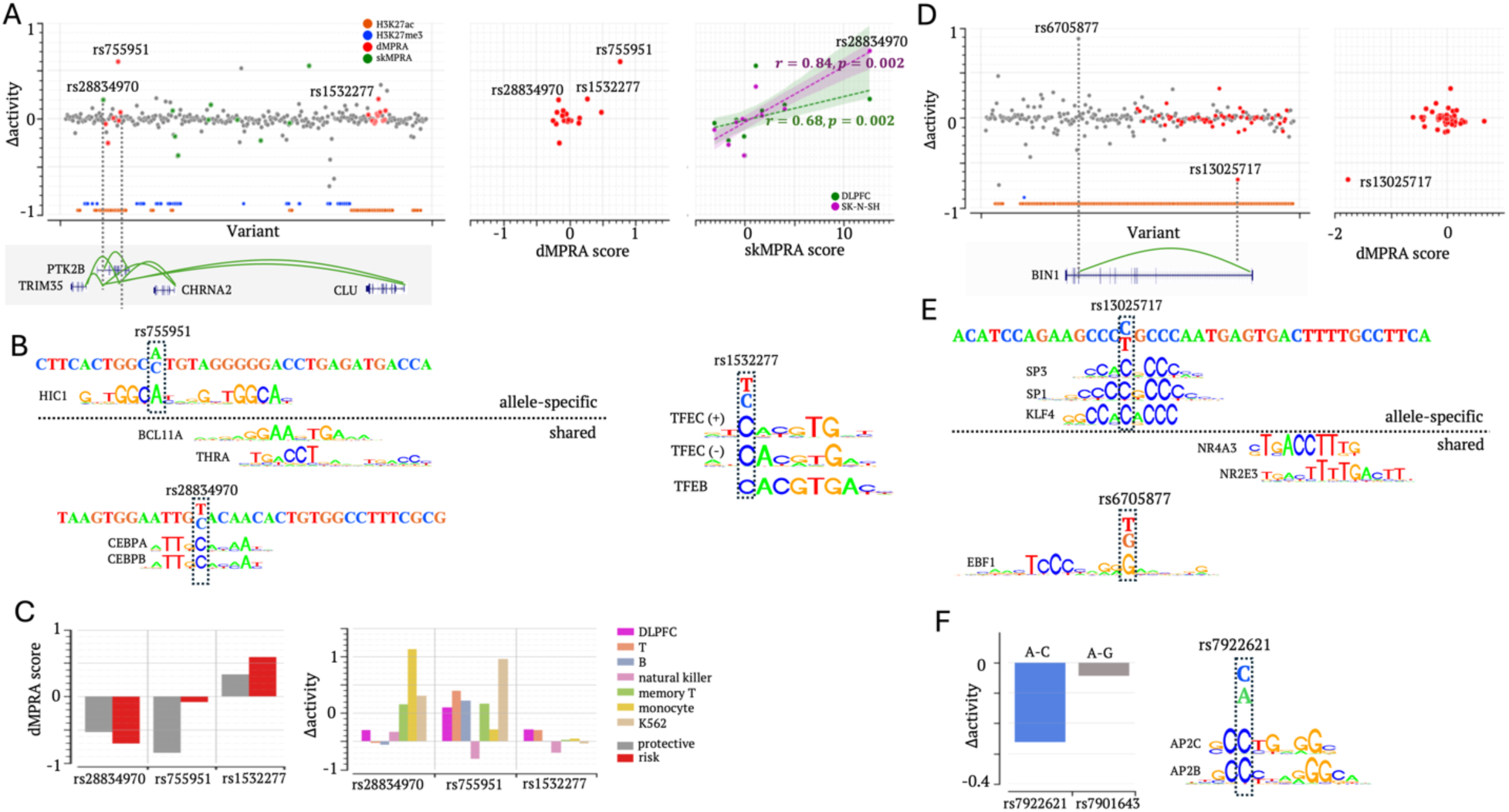
Prioritizing radSNVs in additional AD susceptibility loci. (A) Δactivity and dMPRA scores in the *PTK2B-CLU* locus. Left: Δactivity scores; middle and right: comparisons of Δactivity scores with dMPRA and skMPRA results, respectively. (B) TF motif mapping analysis for rs755951, rs28834970, and rs1532277. (C) dMPRA results (the left panel) and Δactivity scores (the right panel) for rs755951, rs28834970 and rs1532277. (D) Δactivity and dMPRA scores in the *BIN1* locus. (E) TF motif mapping analysis for rs13025717 and rs6705877. (F) Δactivity scores for rs7922621 and rs7901634 in the DLPFC.

Another notable radSNV, rs28834970, resides in an intronic enhancer of *PTK2B* in neuronal cells. This variant is associated with increased *PTK2B* expression in blood cells and an elevated abundance of phosphorylated tau in cerebrospinal fluid ^74^. Although insignificant in dMPRA, this variant shows significant positive Δactivity scores in the DLPFC and SK-N-SH cells, supported by a significant positive skMPRA score (Figure 8A). In blood cells (such as monocytes), the T-to-C substitution at this variant has significantly positive Δactivity scores (Figure 8C), consistent with increased *PTK2B* expression in blood cells (Figure S14). These positive Δactivity scores are further supported by the finding that the risk allele T introduces binding motifs of the activator CEBP family (Figure 8B). All these validate Δactivity predictions at this variant in the DLPFC and blood cells. Interestingly, in this locus, SK-N-SH Δactivity scores show a stronger correlation with skMPRA (*r* = 0.84, *P* = 0.0002) than the DLPFC Δactivity scores (*r* = 0.68, *P* = 0.0002, the third panel in Figure 8A), showcasing the ability of TREDNet models to capture cell-specific regulatory signatures.

Additional examples of identified causal radSNVs are in the *BIN1* locus, an EN locus. Among 51 dMPRA-tested radSNVs, rs13025717 shows the lowest Δactivity, coinciding with its lowest dMPRA score (Figure 8E). The C-to-T substitution at this SNV reduces the ATAC-seq signal and disrupts the binding motifs for SP1 and KLF4, a transcriptional activator and partner of PU.1 (Figure 8F) ^65^. These together validate the significant negative Δactivity score. Another identified AD causal variant in this locus is rs6705877, an intronic SNV not tested in MPRA experiments. The risk allele T at this variant generates a binding motif for EBF1 (Figure 8E) and is associated with increased *BIN1* expression, as reported in the GTEx data (Figure S14). With these findings, rs6705877, carrying the most significant Δactivity in the *BIN1* locus, is a strong candidate for further investigation.

In the *TSPAN14-MAT1A* locus, two proximal SNVs – rs7922621 and rs7910643, located 225 bp apart – reside within the same intronic enhancer ^62^. Genome editing experiments confirmed rs7922621, rather than rs7910643, as the causal variant for the downregulation of *TSPAN14* ^62^. These results are accurately reflected by Δactivity = -0.262 and -0.042 for rs7922621 and rs7910643, respectively. The C-to-A substitution at rs7922621 disrupts binding motifs for AP2C and AP2B (Figure 8F), providing mechanistic insights into its regulatory effect.

## Discussion

The escalating global prevalence of AD, coupled with its pronounced heritability, has spurred decades of research into the genetic architecture of this disease. Despite significant advancements, inadequate annotation of regulatory SNVs associated with AD (adSNVs) remains a major obstacle to progress. To address this gap, we developed a deep learning framework, TREDNet, to predict the regulatory impact of these variants. Using this approach, we detected 1,457 silencer and 3,084 enhancer AD variants (collectively termed radSNVs) in DLPFC. radSNVs show stronger GWAS AD associations compared to the general pool of adSNVs. Silencer radSNVs are predominantly associated, via proximity or eQTLs, with genes lowly expressed in the healthy DLPFC, whereas enhancer radSNVs are associated with highly expressed genes.

Assessing the regulatory influence of radSNVs is essential for elucidating genetic underpinnings of AD. Nucleotide-resolution tools such as MPRA assays and machine learning models are instrumental in this pursuit, offering nucleotide-level insights into regulatory mechanisms ^19–21,76,77^. However, as prior studies have noted, MPRA assays often exhibit biases. Their low sensitivity of experimental constructs and signal-calling strategies to silencing sequences result in an overrepresentation of SNVs with at least one activating allele ^63,78^. For example, in skMPRA, 13,566 variants carry H3K27ac signals in the DLPFC, while only 3,731 variants show H3K27me3 modification. Similarly, in dMPRA, 1612 and 207 variants overlap with H3K27ac and H3K27me ChIP-seq peaks in the DLPFC, respectively. In this context, machine learning models present a promising alternative. Our TREDNet model, which explicitly quantifies silencer and enhancer activity by analyzing DNA sequence composition, demonstrates robust predictive power of silencers and enhancers, as well as their disruptive variants, as evidenced by significant correlations with experimental results from MPRAs and beyond. Applying this framework, we identified causal radSNVs for AD. These variants are characterized by notable enrichment in neuronal cell TF ChIP-seq peaks, brain-specific TF binding motifs, and evolutionarily conserved regions, confirming their pivotal regulatory roles.

The distribution of radSNVs categorizes AD susceptibility loci into three classes – 14 SL loci, 44 EN loci, and 13 ENSL loci. Each class, exhibiting unique transcriptomic and epigenomic patterns during AD progression, is associated with specific biological functions, molecular activities, and perturbation responses. For example, genes associated with SL loci are suppressed in the healthy DLPFC but upregulated in the AD DLPFC, accompanied by reduced H3K27me3 and increased H3K27ac intensities in AD patients. EN-locus genes often govern metabolic processes, such as gated channel activities and lipid tube assembly, and are modulated by channel blockers such as BAPTA-AM and quinidine. ENSL loci are chiefly associated with tau-related pathways and synapse degeneration. In contrast, SL loci are pivotal for neuroimmune response and microglia regulation. Moreover, each locus class has unique phenotypic associations. SL radSNVs are genetic components frequently shared by AD and autoimmune disorders; ENSL radSNVs are common genetic factors for both AD and tau-associated neurological diseases, such as PD; EN loci are exclusively associated with Lewy bodies. Collectively, by categorizing AD susceptibility loci, we delineate the complex regulatory landscape of AD into distinct biological pathways, providing regulatory insights for its pathogenesis.

Our silencer and enhancer profiles were also applied to illuminate molecular and cellular features of five main AD subtypes. Using them, we reported that A*β*-overaccumulation subtypes are characterized by SL-locus gene upregulation, overreactive microglia and astrocytes, and excitatory neuron loss. Tau-upregulation subtypes are marked by upregulated ENSL-locus genes and hyperactive excitatory neurons. Many of these findings align with experimental observations, validating the contribution of our framework and providing new avenues for mechanistic interpretation of AD phenotypes across different patient cohorts.

Further analysis of SL loci highlights their significant and unique role in the dysregulation of AD microglia. Over half (54.2%) of SL-locus genes are upregulated in AD microglia, representing a four-fold enrichment compared to the genome-wide average. Moreover, 70% of SL-locus radSNV eQTLs are microglia-specific, far exceeding the 39% observed for all brain eQTLs. Strikingly, 90% of these eQTLs have AD risk alleles associated with gene upregulation, representing a significant enrichment compared to all microglia eQTLs. SL-locus genes are often induced by both INF-γ and pre-formed Aβ fibrils in iMGL cells. These findings underscore SL-locus radSNVs as a core modulator for aberrant microglial response, a defining feature of AD pathology ^79^. We further pinpoint the potential causal radSNVs for this abnormality. For example, the silencer SNVs rs667897 and rs636317, which disrupt the binding motifs of CTCF and NFE, are likely drivers for the dysregulation of *MS4A* genes in the AD DLPFC.

As a final note, our deep learning model can be further extended to include additional functional AD data. For example, by adding spatial single-cell ATAC-seq data, our model can mark silencers and enhancers for different DLPFC (or brain) sections, facilitating the investigations into brain spatial-specific attributes for AD subtypes.

In summary, by mapping enhancers and silencers in the DLPFC, this work advances the understanding of the molecular basis of AD. Despite being only half as abundant as enhancers radSNVs, silencer radSNVs exert substantial and unique influences on AD pathogenesis, particularly through their unique source of variance and distinguishing phenotypic impact on SL loci. As silencer variants have also been implicated in different diseases, such as breast cancer ^80^, Schizophrenia ^81^, and facial paresis ^82^, yet remain largely underexplored, our work presents a framework of silencer variant analysis, which can be readily extended to the other diseases. Systematic investigation of silencer variants is essential for a comprehensive understanding of the genetic architecture underlying complex diseases like AD.

## Methods

### Predicting silencer and enhancer variants in the DLPFC using a modified TREDNet

We modified the TREDNet model, a two-phase deep learning model ^25^, to predict enhancers and silencers in the DLPFC and eight major cell types. We downloaded DNase-seq peaks, H3K27ac, and H3K27me3 ChIP-seq peaks (“narrow peak”) in the DLPFCs of 20 undemented postmortem brains cataloged in the RUSH project (https://www.encodeproject.org/brainmatrix/?type=Experiment&status= released&internal_tags=RushAD). For each sample, candidate enhancers were defined as the DNase-seq peaks overlapping H3K27ac ChIP-seq peaks but not H3K27me3 peaks in the central 400bp. Candidate silencers were the DNase-seq peaks overlapping H3K27me3 peaks but not H3K27ac peaks in the central 400bp, together with the non-DNase-seq-overlapping H3K27me3 peaks not overlapping H3K27ac peaks. Each of these elements was then extended to a 1kbp-long sequence centering at its midpoint. To ensure the specificity for silencers and enhancers, the elements overlapping with promoters and exons of protein-coding genes annotated in the GENCODE project ^83^ were excluded. After merging data from different samples, redundancy across elements was addressed by randomly retaining an element among those overlapping with each other by more than 600bp.

To annotate the cell-specific functions of candidate elements, we integrated single-cell ATAC-seq data from three studies ^17,27,28^, encompassing chromatin accessibility profiles for 8 brain cell types, in our deep learning model. To this end, our model profiles a set of predicted enhancers and silencers for nine cellular contexts: the DLPFC and eight distinct brain cell types. Control samples were generated by randomly sampling DNase-seq peaks documented for non-brain biosamples in the ENCODE project ^29^, excluding those overlapping with candidate silencers and enhancers, as well as epigenomic peaks used to define these elements.

To predict enhancers and silencers for 9 cellular contexts, the TREDNet model consists of 19 output nodes with the activation function of “SoftMax”, corresponding to silencers and enhancers for nine cellular contexts, as well as controls. For each cellular context, the cost function was the “categorical cross entropy” of three classes – enhancer, silencer, and control. The overall cost function for training this 19-output TREDNet model was the sum of these context-specific “categorical cross entropy” functions.

We collected AD-associated variants from three studies ^14,23,24^. The AD association of a SNV is the minimum of AD association p-values across these studies. SNVs are considered as AD-associated if their AD-associations are less than 10^−5^, dubbed as adSNVs. To evaluate the regulatory activities of a non-coding adSNV, we used the 1kbp sequence centering at this variant as the input to the TREDNet model. The outputs of the TREDNet model are our prediction scores of regulatory activities of input variants. A variant was labelled as a silencer variant if it was located within a H3K27me3 peak detected in the DLPFC (as documented in the ENCODE RUSH project) and its silencer prediction score exceeded the threshold *C*_*s*,*i*_ in at least one of cellular contexts (where *i* represents a cell context). In a cellular context *i*, *C*_*s*,*i*_ was the silencer prediction score corresponding to a false positive rate (FPR) of 0.05 among test samples wherein 90% were controls. Similarly, a variant was predicted as an enhancer variant if its enhancer prediction score *y*_*e*_ was greater than the cutoff *C*_*e*_ in at least one cellular context and it resided within a H3K27ac peak in the DLPFC. In a cellular context *i*, *C*_*e*,*i*_was the prediction score corresponding to the FPR of 0.05 among a test sample set in which 90% were controls.

### Δactivity of regulatory variants

As detailed in our prior study ^63^, the regulatory difference between two alleles for a given variant is measured as

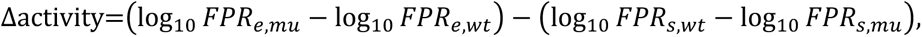

where *wt* and *mu* represent the wild type and mutant alleles, respectively. *FPR*_*e*_ is the false positive rate for enhancer prediction scores among a sample set comprising 90% controls. Similarly, *FPR*_*s*_ is the false positive rate for silencer prediction scores. A positive Δactivity indicates a mutation-driven increase in activation impact potential.

### Categorization of AD susceptibility loci

We integrated AD-associated variants from three recent GWAS studies ^14,23,24^, resulting in a set of 18,826 adSNVs distributed across 1475 gene loci. A gene locus encompasses the gene body and flanking upstream and downstream intergenic regions. After merging adjacent gene loci hosting adSNVs, we retrieved 124 genomic regions enriched with adSNVs compared to the genome-wide average (*P* < 0.003, Table S1). These regions are referred to as AD susceptibility loci. As such, each locus probably spans multiple gene loci. We used gene annotations cataloged in the GENCODE project ^83^ to categorize AD-associated variants into two broad classes: promoter/exon and distal-RE variants.

To evaluate the enrichment of silencer radSNVs within an AD susceptibility locus (say *j*), we compared their density to that across the entire genome:

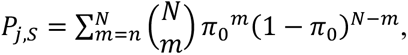

where *π*_0_ is the ratio of the locus length to the human genome size. *N* is the total number of silencer radSNVs, and *n* is the number of silencer radSNVs in the locus *j*. The locus *j* was defined as enriched for silencer radSNVs when *P_j_*_,*S*_ < 0.005. Similarly, the locus *j* is enriched for enhancer radSNVs when *P_j_*_,*E*_ < 0.005. Loci enriched exclusively for silencer radSNVs were classified as silencer (SL) loci, those enriched exclusively for enhancer radSNVs as enhancer (EN) loci, and those enriched for both as enhancer-silencer (ENSL) loci.

### Enrichment of GWAS SNVs among radSNVs in individual loci

To investigate the overlap of radSNVs with GWAS SNVs associated with a disease (*d*) in a given locus (*j*), we used the genome-wide expectation as the control. That is,

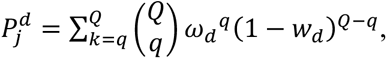

where *Q* represents the number of radSNVs in the locus *j*, and *q* is the number of these radSNVs associated with the disease *d*. Here, *w*_d_ denotes the fraction of the common variants associated with the disease *d*. Enrichment was considered significant if 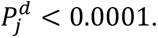

### Differential signal analysis for H3K27ac and H3K27me3 modification between healthy and AD DLPFCs

H3K27ac and H3K27me3 ChIP-seq data (narrow peak and bam files) were downloaded from the RUSH project (https://www.encodeproject.org/brainmatrix/?type=Experiment&status=released&internal_tags=RushAD). We merged narrow ChIP-seq peaks as histone modification regions and then utilized DESeq2 ^84^ for evaluate epigenetic signal difference between healthy and AD cases in these merged peaks. Using *P* < 0.005, 4499 H3K27ac peaks and 2799 H3K27me3 peaks were identified to exhibit significant intensity difference (Table S3). To address long-distance chromatin interactions, a histone modification region was associated with an AD susceptibility locus within 1Mpb to its center.

### Predicting gene expression in AD subtypes using linear regression

To investigate the associations between AD subtype and brain cell types, we utilized a linear regression model to predict AD-subtype gene expression using enhancer/silencer and gene profiles of seven brain cell types. Differential gene expression profiles between healthy and AD brains were sourced from publicly available datasets, i.e., the ssREDA resource ^60^ and a dataset from the study of S. Morabito et al. ^27^. Enhancer/silencer profiles were established by applying the TREDNet model to single-cell ATAC-seq peaks from three recent studies ^17,27,28^. ATAC-seq peaks were extended to 1kb at their midpoints and used as the TREDNet input. The regulatory influence of an ATAC-seq peak (say *i*) in a cell type 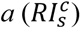 is measured as

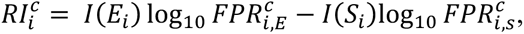

where 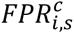 is the false positive rate for its silencer prediction score and 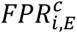 is the false positive rate for its enhancer prediction score. *I*(*E*_*i*_) = 1 if the ATAC-seq peak *i* overlap with a H3K27ac peak in the DLPFC and *I*(*E*_*i*_) = 0 otherwise. *I*(*S*_*i*_) = 1 if the ATAC-seq peak *i* overlap with a H3K27me3 peak in the DLPFC and *I*(*S*_*i*_) = 0 otherwise.

Regulatory domains for a gene include the genomic regions surrounding its TSS with 200kbp and its whole intronic region. In the cell type *c*, the regulatory impact on this gene was quantified as the sum of 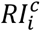 among ATAC-seq peaks located within its regulatory domain. Furthermore, to normalize contribution across cell types and data types, gene expression and regulation impact values were scaled to a maximum of 1 within each cell type and data type.

We applied the Lasso linear model (from scikit-learn library, with the setting of alpha =0.0001, precompute=True, max_iter=1000 and selection=‘random’) to predict AD subtype gene expression using three input types: (1) cell-type gene expression profiles, (2) cell-type regulatory impact profiles, and (3) a combination of both. Model performance was evaluated under a ten-fold validation, averaging RMSEs across 100 trails for each input type (Figure 5D). Model weights, which can be interpreted as the correlations between cell types and AD subtypes, were derived as the average weights across all built models. For the models trained on the combination of gene expression and enhancer/silencer profiles, the weights are the average of the weights from different data types.

### Publicly available data and tools used in this study

The Genotype-Tissue Expression Project used here is GTEx database version 8 ^35^, documenting eQTLs for 13 brain regions and the whole blood. All brain eQTLs were merged into an eQTL set for brain tissue. An eQTL (i.e., a variant and its associated gene) was considered tissue-specific if it was detected only in a single tissue. eQTLs for brain cell types were downloaded from the study of M. Fujita et al. ^36^ Similarly, a brain eQTL was considered cell-specific if it was only detected in a single brain cell. Blood DICE eQTLs ^58^ were sourced at https://dice-database.org/downloads. To ensure high cell-specificity of DICE eQTLs, we clustered DICE blood cell types based on the similarities of their eQTL profiles, resulting in five groups of DICE cell types (see Supplementary Notes). A DICE eQTL was considered cell-specific if it was only detected in a single cell-type group.

We collected 1) GWAS SNVs curated in the NHGRI catalog and in UK Biobank release 2 cohort and 2) the SNVs in tight linkage disequilibrium (LD r^2^ > 0.8) to these SNVs based on EUR population in 1000 Genomes Project ^63^ as the control for analyzing the association of radSNVs with other disorders and the enrichment of radSNVs among eQTL SNVs (Figures 3C, E, and G). Here, using GWAS SNVs, rather than all common SNVs, is to address the potential distribution bias of SNVs examined in GWAS and eQTL studies.

TF ChIP-seq data for SK-N-SH cells were downloaded from the ENCODE project. TF binding motifs were retrieved from the MEME Suite (https://meme-suite.org/meme/db/motifs). The Find Individual Motif Occurrence (FIMO), with the default setting, was used to map motifs in genomic sequences with the default setting. The regulatory effect of a binding motif was quantified by comparing its overlap with H3K27ac ChIP-seq peaks in the healthy DLPFC with that with H3K27me3 peaks. That is, for a motif (say, *m*), we have

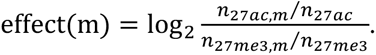

Here, *n*_27*aC*_ represents the numbers of adSNVs carrying H3K27ac ChIP-seq signals, while *n*_27*aC*,*m*_ is the count of these H3K27ac adSNVs overlapping with the motif *m*. Given that H3K27ac and H3K27me3 modifications are associated with transcriptional activation and suppression, a positive/negative effect(m) denotes an activating/repressive binding motif.

We downloaded the 100-way evolutionarily conserved elements from the UCSC genome data resource at https://hgdownload.soe.ucsc.edu/goldenPath/hg38/database/. Cell-specific compound-responsive gene lists were from the L1000 project at https://maayanlab.cloud/sigcom-lincs/#/Download. H3K27ac HiChIP chromatin contact data were retrieved from the HiChIPdb ^38^ at https://health.tsinghua.edu.cn/hichipdb/about.php. DESEQ2 were used to detect the H3K27ac and H3K27me3 ChIP-seq peaks having significantly different signals between healthy and AZ DLPFCs.

## Data Availability

The built model, data, and codes are available at https://zenodo.org/records/15127846.

## Data and code availability

The built model and codes are available at https://zenodo.org/records/15127846. Samples used for training the TREDnet model is available at https://zenodo.org/records/15127846.

## Competing interests

The authors declare that they have no competing interests.

## Acknowledgments

This research was supported by the Division of Intramural Research of the National Library of Medicine. In addition, this work utilized the computational resources of the NIH HPC Biowulf cluster (http://hpc.nih.gov).

## Notes

### Competing Interest Statement

The authors have declared no competing interest.

