## Supplementary material for "Silencer variants are key drivers of gene upregulation in Alzheimer’s disease": supplmentary notes

Di Huang and Ivan Ovcharenko*

Division of Intramural Research, National Library of Medicine, National Institutes of Health, Bethesda, MD, 20892, USA.

### Supplementary Notes

#### Tissue-specificity of genes based on gene expression profiles.

We downloaded the RNA-seq data of 251 biosamples from the ENCODE project. The gene expression, measured as the Reads Per Kilobase of the transcript, per Million mapped reads (RPKM values), were normalized so that the expression level of each gene had a median of zero. Tissue-specificity of a gene was measured using tau (τ) (1) as

$\tau=\frac{\sum_{i=1}^{N} (1-\hat{x}_{i})}{N}, \hat{x}_{i}=\frac{x_{i}}{{max}_{k=1}^{N}x_{k}}$,

where $x_{i}$ is the expression of a tested gene in the cell line $i$. $N$ is the number of the cell lines under consideration. The values of $\tau$ are in the range of $[0,1]$. A high value of $\tau$ corresponds to a large variation in gene expression across tissues, i.e., a high tissue specificity. The genes with $\tau>0.97$ were considered as tissue-specific, while the genes with $\tau<0.8$ were labelled as housekeeping.

### Supplementary Figures

#### Supplementary Fig. S1

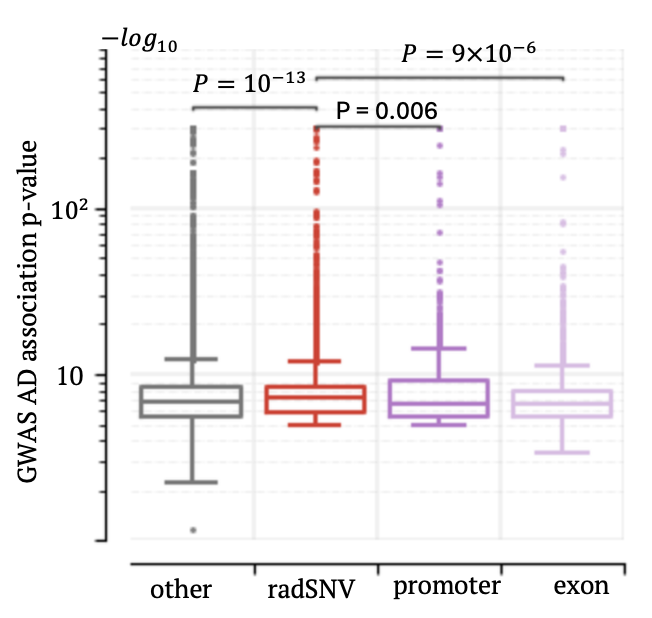

**Figure S1.** GWAS association significance with AD ($-\log_{10} P$) across adSNVs groups. The AD-association $p$-value of a variant is the smallest association $p$-value across the examined three AD GWAS studies.

#### Supplementary Fig. S2

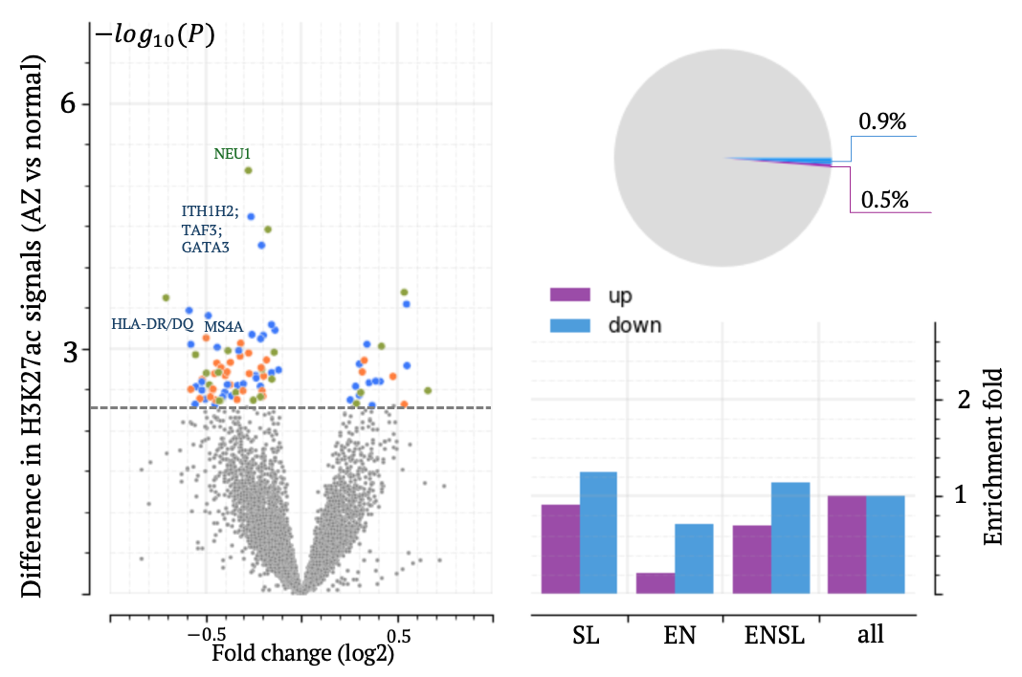

**Figure S2.** Changes in H3K27me3 signals between healthy and AD DLPFCs (the volcano plot in the left panel), and their distribution across different locus classes (the bar plot and the pie chart in the right panel). In the volcano plot, each dot represents a H3K27me3 peak. Grey dots indicate a peak with an insignificant change. Blue, orange, and blue dots indicate peaks with significant changes located in SL, EN, and ENSL loci, respectively.

#### Supplementary Fig. S3

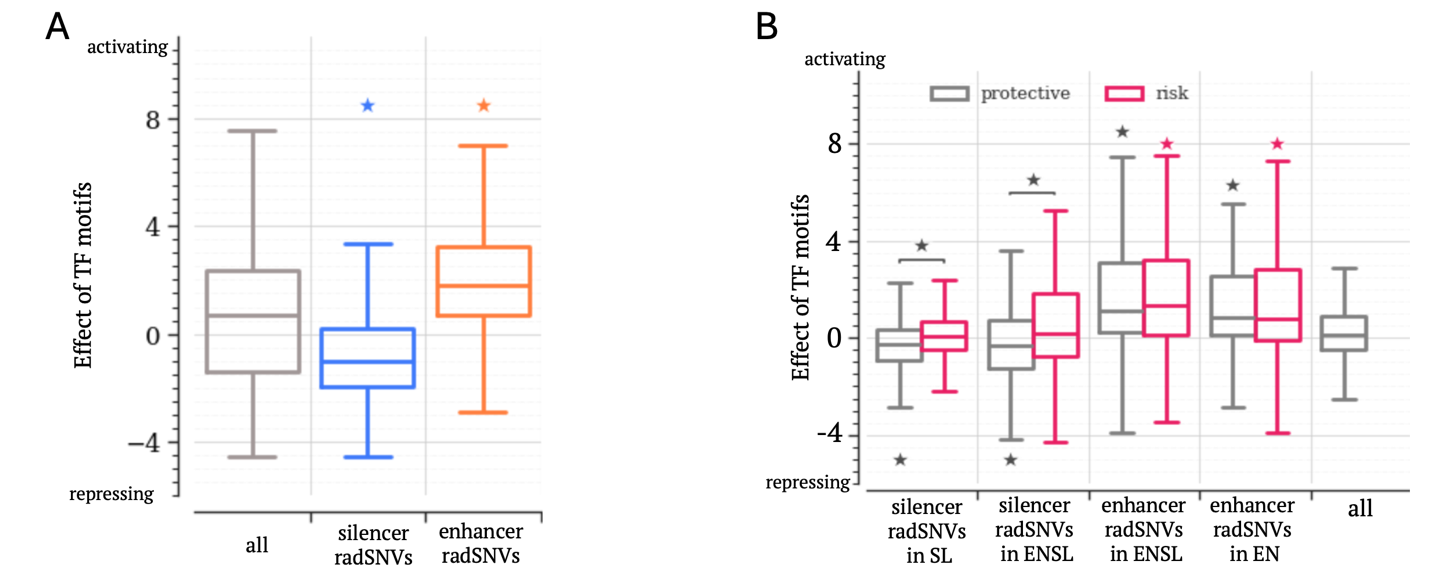

**Figure S3.** Effect scores of TF motifs enriched among different radSNV groups: (A) among silencer and enhancer radSNVs with protective alleles; (B) among silencer and enhancer radSVNs in different locus classes for protective and risk alleles. Controls for motif enrichment analysis are all regulatory adSNVs. Asterisks above bars indicate the significant difference ($p<{10}^{-5}$) in effect scores from the motifs enriched in either silencer or enhancer radSNVs (denoted as “all” in the plots). The upper and lower whisker edges in these boxplots represent approximately 25% and 75% quartiles of the presented data.

#### Supplementary Fig. S4

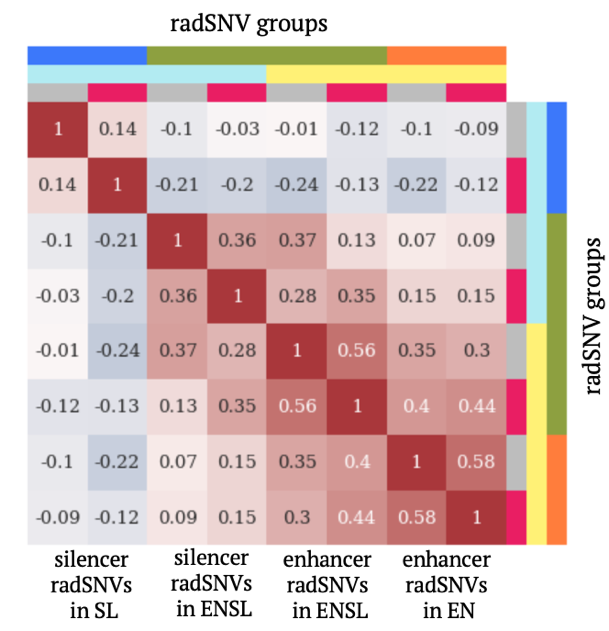

**Figure S4.** Similarity among TF Motif profiles across radSNV classes for protective and risk alleles.

#### Supplementary Fig. S5

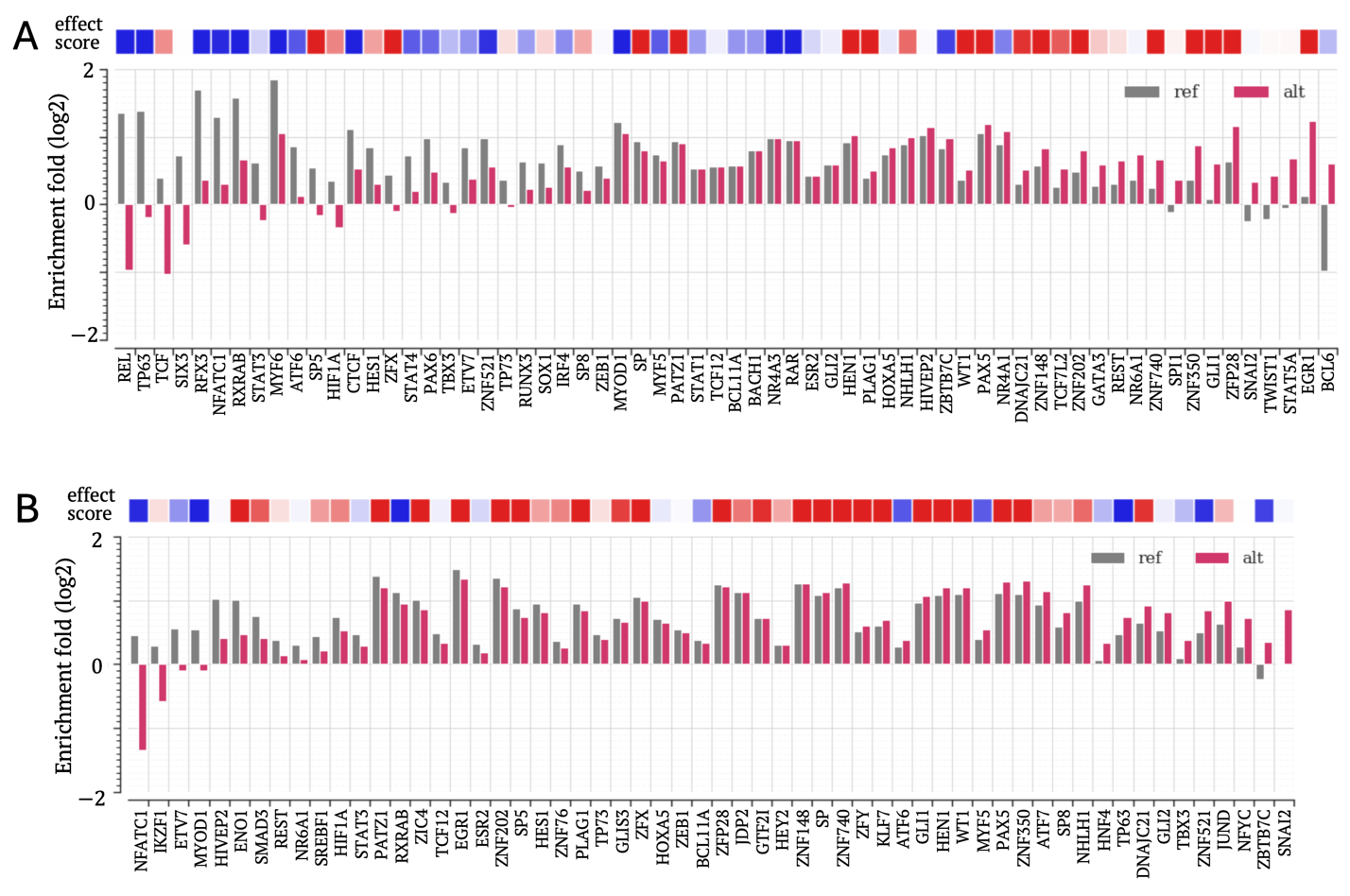

**Figure S5.** TF Motifs enriched for protective and risk alleles among (A) silencer and (B) enhancer radSNVs. Controls for motif enrichment analysis are all regulatory adSNVs.

#### Supplementary Fig. S6

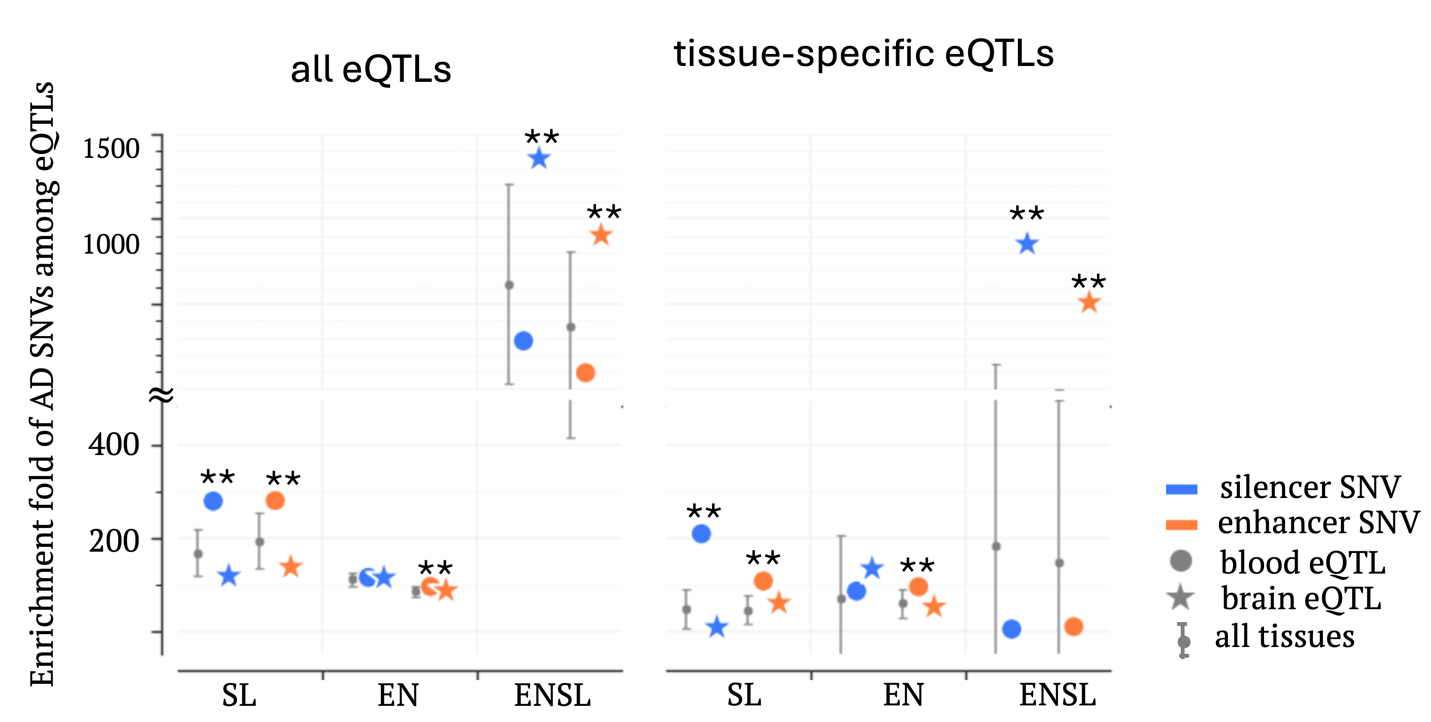

**Figure S6.** Enrichment of radSNVs among eQTLs in brain and blood across locus classes. RadSNVs of a locus class were stratified into two groups: silencer SNVs and enhancer SNVs. The results demonstrate the comparable regulatory importance between silencer and enhancer SNVs. The upper and lower whisker edges in these bars represent approximately 25% and 75% quartiles of the presented data. $**:P<{10}^{-5}$.

#### Supplementary Fig S7

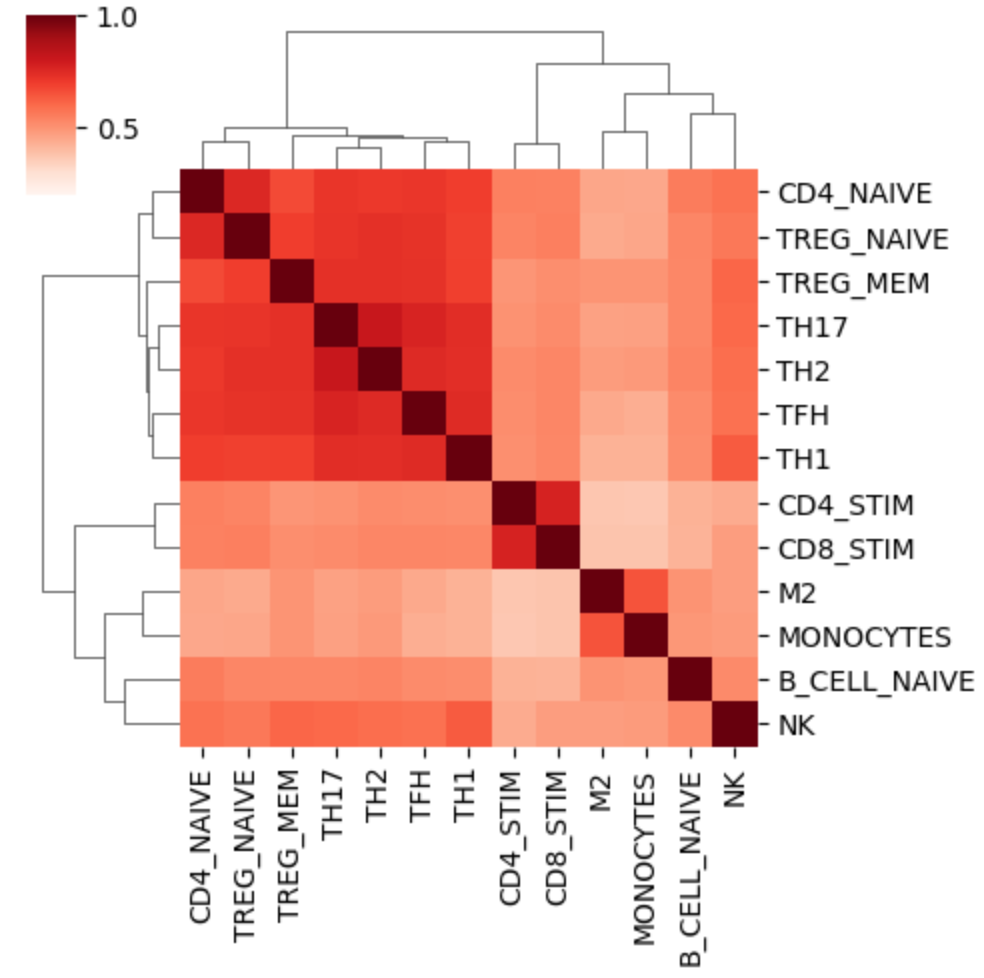

**Figure S7.** Similarities of DICE eQTLs across blood cell types. Based on these similarities, tested blood cell types are clustered in four groups. Jaccard index was used to measure the similarities (i.e., the overlaps) between eQTL sets of blood cell types.

#### Supplementary Fig. S8

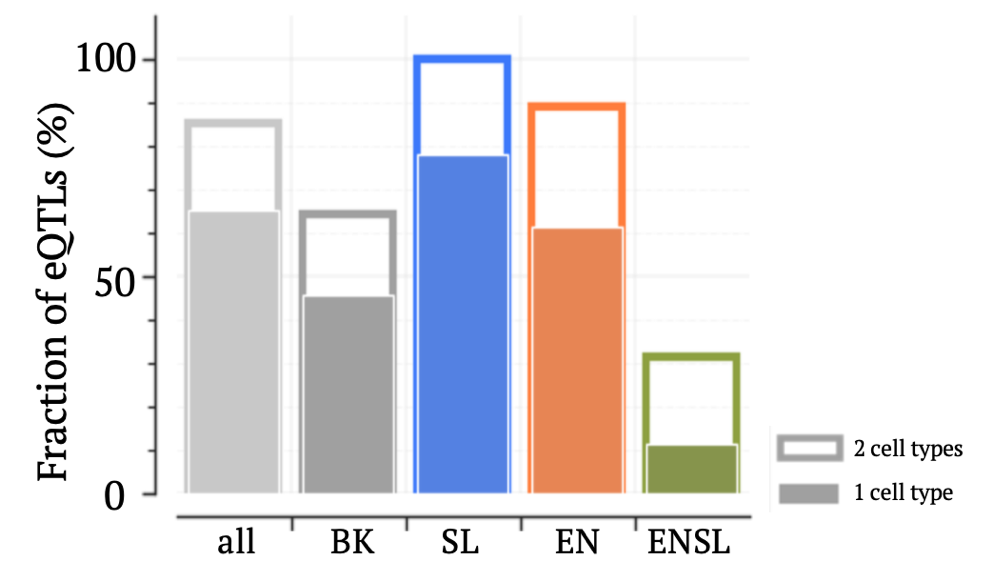

**Figure S8.** Cell-specificity of brain eQTLs across locus classes. Cell-specificity of an eQTL is measured as the number of cell types where that eQTL is active.

#### Supplementary Fig. S9

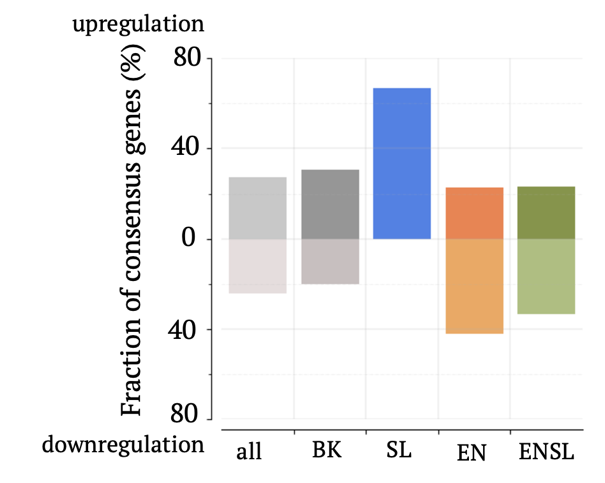

**Figure S9.** Fraction of INF-γ-up/down-regulated genes that are induced/repressed by pre-formed Aβ fibrils across locus classes.

#### Supplementary Fig. S10

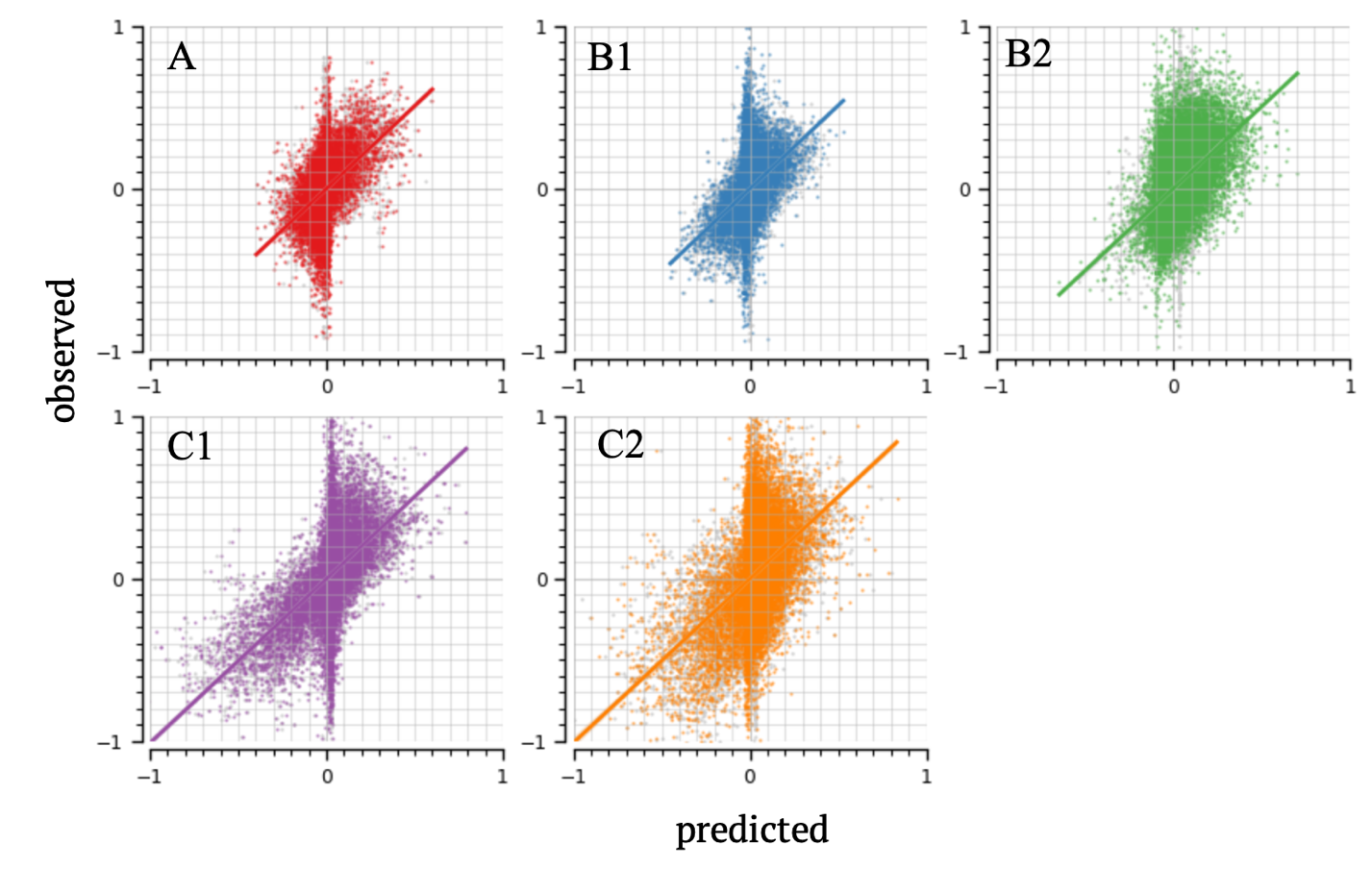

**Figure S10.** Comparison of observed and predicted gene expression across AZ subtypes.

#### Supplementary Fig. S11

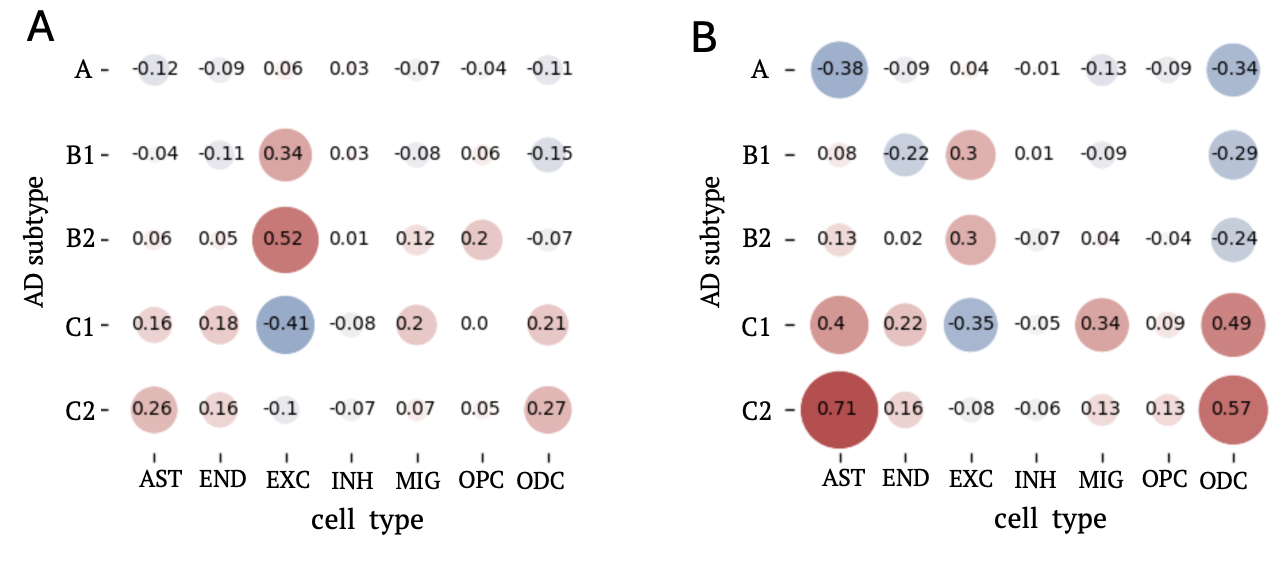

**Figure S11.** Average weights in linear regression models built on (A) cell-specific gene expression data and (B) enhancer/silencer profiles.

#### Supplementary Fig. S12

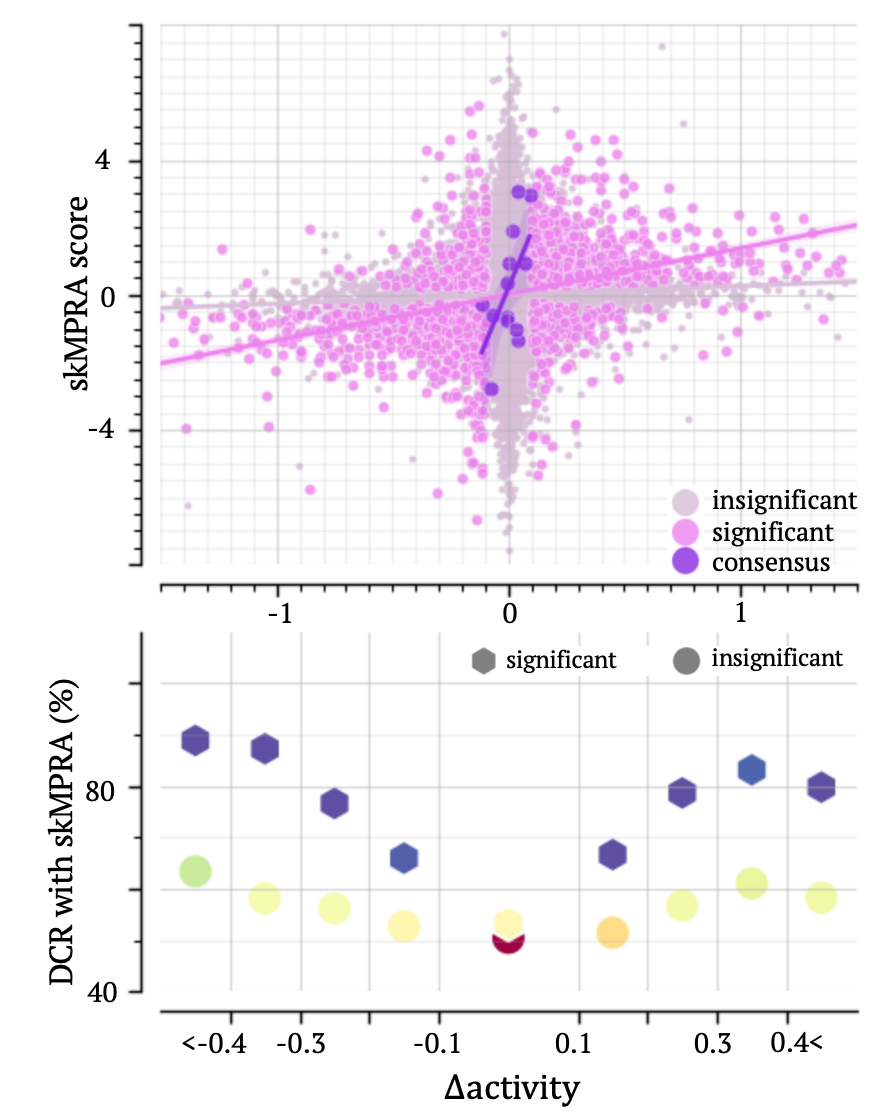

**Figure S12.** Comparison between skMPRA and $\text{∆activity}$scores predicted by the SK-H-SH TREDNet model built in our prior study (2). (A) Correlation between $\text{∆activity}$ and skMPRA scores. (B) DCRs between skMPRA and $\text{∆activity}$ scores for variants, stratified by $\text{∆activity}$.

#### Supplementary Fig. S13

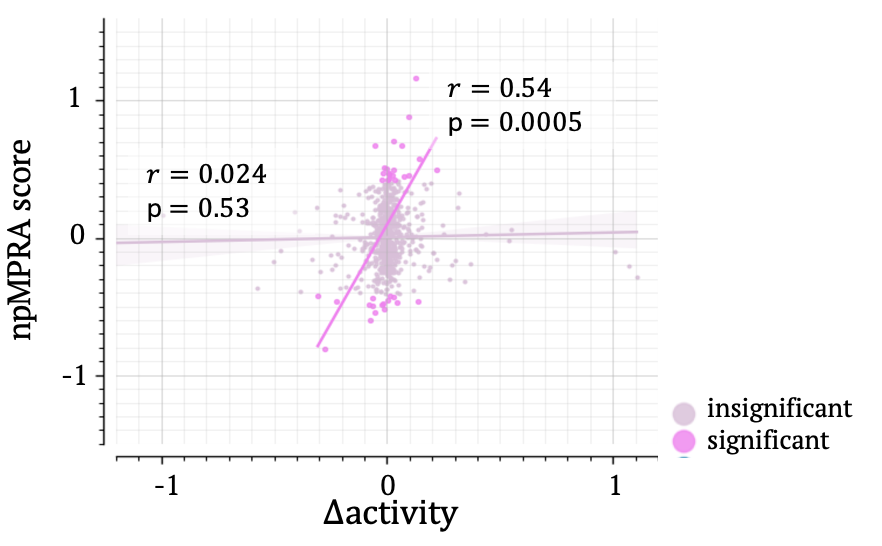

**Figure S12.** Comparison between npMPRA and $\text{∆activity}$scores in DLPFC.

#### Supplementary Fig. S14

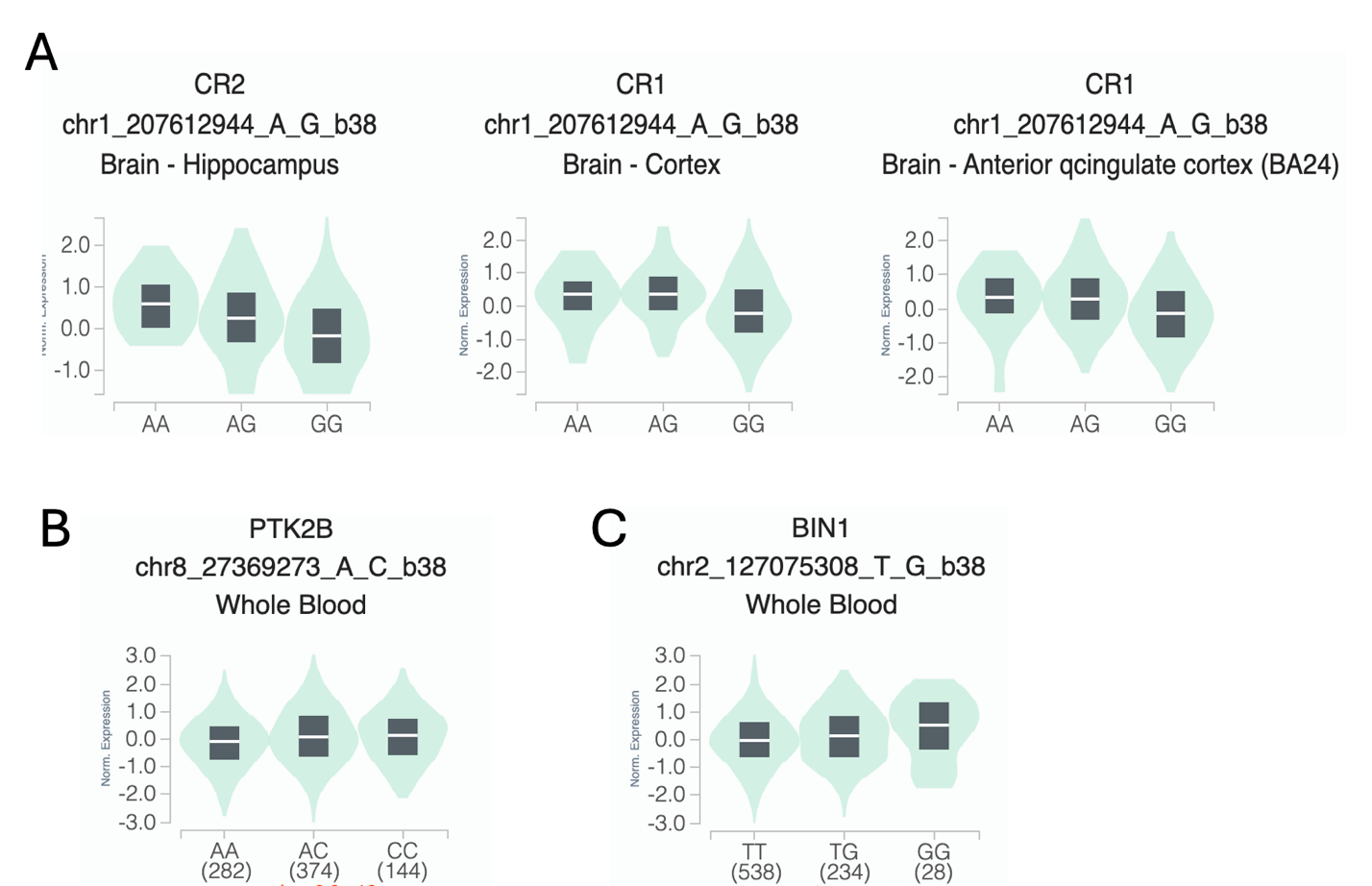

**Figure S14.** GTEx results for eQTLs (A) rs6701713 (B) rs755951 and (C) rs6705877.

#### Supplementary Fig. S15

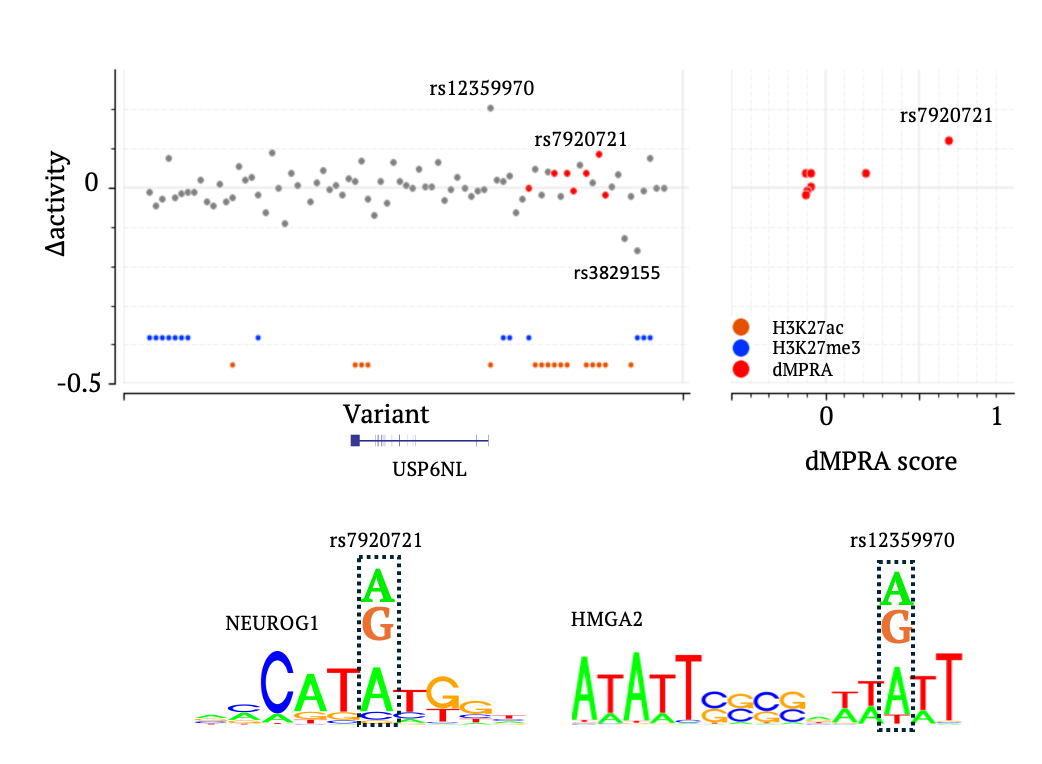

**Figure S15.** Prioritizing radSNVs in the *USP6NL* locus.

**Table S1.** List of AD susceptibility loci.

**Table S2.** List of regulatory AD-associated SNVs (radSNVs). Predicted $\Delta\text{activity}$ scores of these variants are provided.

**Table S3.** H3K27ac and H3K27me3 peaks having significantly different intensities between healthy and AD cases.

1.	Kryuchkova-Mostacci N, Robinson-Rechavi M. A benchmark of gene expression tissue-specificity metrics. Briefings in Bioinformatics. 2017;18(2):205-14.

2.	Huang D, Ovcharenko I. The contribution of silencer variants to human diseases. Genome Biology. 2024;25(1):184.
